# Maternal smoking DNA methylation risk score associated with health outcomes in offspring of European and South Asian ancestry

**DOI:** 10.1101/2023.09.24.23295907

**Authors:** Wei Q. Deng, Nathan Cawte, Natalie Campbell, Sandi M. Azab, Russell J de Souza, Amel Lamri, Katherine M. Morrison, Stephanie A. Atkinson, Padmaja Subbarao, Stuart E. Turvey, Theo J. Moraes, Koon K. Teo, Piush Mandhane, Meghan B. Azad, Elinor Simons, Guillaume Pare, Sonia S. Anand

## Abstract

Maternal smoking has been linked to adverse health outcomes in newborns but the extent to which it impacts newborn health has not been quantified through an aggregated cord blood DNA methylation (DNAm) score. Here we examine the feasibility of using cord blood DNAm scores leveraging large external studies as discovery samples to capture the epigenetic signature of maternal smoking and its influence on newborns in White European and South Asian populations. We first examined association between individual CpGs and cigarette smoking during pregnancy, smoking exposure in two White European birth cohorts (n = 744). Several previously reported genes for maternal smoking were supported, with the strongest and most consistent signal from the *GFI1* gene (6 CpGs with *p* < 5×10^-5^). Leveraging established CpGs for maternal smoking, we constructed a cord blood epigenetic score of maternal smoking that was validated in one of the European-origin cohorts (n = 347). This score was then tested for association with smoking status, secondary smoking exposure during pregnancy, and health outcomes in offspring measured after birth in an independent white European (n = 397) and a South Asian birth cohort (n = 504). The epigenetic maternal smoking score was strongly associated with smoking status during pregnancy (OR=1.09 [1.07,1.10], *p*=5.5×10^-33^) and more hours of self-reported smoking exposure per week (1.93 [1.27, 2.58], *p*=7.8×10^-9^) in White Europeans, but not with self-reported exposure (*p* > 0.05) in South Asians. The same score was consistently associated with a smaller birth size (−0.37±0.12 cm, *p*=0.0023) in the South Asian cohort and a lower birth weight (−0.043±0.013kg, *p*=0.0011) in the combined cohorts. This cord blood epigenetic score can help identify babies exposed to maternal smoking and assess its long-term impact on growth. Notably, these results indicate a consistent association between the DNAm signature of maternal smoking and a small body size and low birthweight in newborns, in both white European mothers who exhibited some amount of smoking and in South Asian mothers who themselves were not active smokers.

## Introduction

Maternal smoking has adverse effects on offspring health including pre-term delivery (1,2), stillbirth (3), and low birth weight (4), and is associated with pregnancy complications such as maternal higher blood pressure, and gestational diabetes (5). Consistent with the Developmental Origins of Health and Disease (DOHaD) hypothesis, maternal smoking exposes the developing fetus to harmful chemicals in tobacco that negatively impact the health of newborns, resulting in early-onset metabolic diseases, such as childhood obesity (6–9). Yet self-reported smoking status is subject to underreporting among pregnant women (10–12). This could subsequently impact the effectiveness of interventions aimed at reducing smoking during pregnancy and may skew data on the risks associated with maternal smoking.

DNA methylation is one of the most commonly studied epigenetic mechanisms by which cells regulate gene expression, and is increasingly recognized for its potential as a biomarker (13). Differential DNA methylation has been established as a reliable biochemical response to cigarette smoking and was shown to capture the long-lasting effects of persistent smoking in ex-smokers (14–16). Recent large epigenome-wide association studies (EWAS) have robustly identified differentially methylated cytosine–phosphate–guanine (CpG) sites associated with adult smoking (15,17,18) and maternal smoking (19,20). Our recent systematic review of 17 cord blood epigenome-wide association studies (EWAS) found that out of the 290 CpG sites reported to be associated with at least one of the following: maternal diabetes, pre-pregnancy body mass index (BMI), diet during pregnancy, smoking, and gestational age, 19 sites were identified in more than one study and all of them associated with maternal smoking (21). Furthermore, these findings have led to a more thorough investigation of the epigenetic mechanisms underlying associations between well-established epidemiological exposures and outcomes, such as the relationship between maternal smoking and birth weight in Europeans (20,22–25) and the less studied African American populations (24) as well as between maternal diet and cardiovascular health (26).

Only a handful of cohort studies were designed to assess the influence of maternal exposures on DNA methylation changes in non-Europeans (24,27). It has been suggested that systematic patterns of methylation (28), such as cell composition, could differ between individuals of different ancestral backgrounds, which could in turn confound the association between differential DNAm and smoking behaviours (29). These systematic differences also contribute to different smoking-related methylation signals at individual CpGs (30). Thus, a comparative study of maternal smoking exposure is a first step towards generalizing existing EWAS results to other populations and a necessary step towards addressing health disparities that exist between populations due to societal privilege, including race or ethnicity and socioeconomic factors.

A promising direction in epigenetic studies of adult smoking is the application of a methylation score (31); this strategy can also be applied to disseminate current knowledge on differential DNA methylation studies of maternal smoking. A methylation score is usually tissue-specific and combines information from multiple CpGs using statistical models (13). Reducing the number of predictors and measurement noise in the data can lead to better statistical power and a more parsimonious instrument for subsequent analyses. It is also of interest to determine whether methylation scores demonstrate the capacity to predict outcomes in diverse human populations, given the presence of systematic differences in methylation patterns due to ancestral backgrounds (28).

In this paper, we investigated the epigenetic signature of maternal smoking on cord blood DNA methylation in newborns, as well as its association with newborn and later life outcomes in one South Asian which refers to people who originate from the Indian subcontinent, and two predominantly European-origin birth cohorts. Similar to the Born in Bradford study (32), we observed several differentiating epidemiological characteristics between South Asian and European-origin mothers. Notably, almost none of the South Asian mothers were current smokers and had low smoking rates pre-pregnancy as compared to European mothers, which is consistent with the broader trends of lower smoking rates in South Asian females (33). Another relevant observation is the small birth size and low birth weight in the South Asian newborns. These differences in newborn size and weight may be influenced by various factors, including maternal nutrition, genetics, and socioeconomic status. Keeping these differences in mind, we first conducted cohort-specific epigenetic association studies between available CpGs and maternal smoking in the predominantly European-origin cohorts, benchmarking with previously identified CpGs for maternal smoking and adult smoking. Second, we leveraged the reported summary statistics from existing large EWASs to construct a methylation risk score (MRS) for maternal smoking. The MRS was first internally validated in one of the European-origin cohorts and then tested in a second independent European-origin cohort. Third, we examined the association between maternal smoking MRS and newborn health outcomes, including length, weight, BMI ponderal index, and early-life anthropometrics in both European and South Asian cohorts.

## Materials and Methods

### Study population

The NutriGen Alliance is a consortium consisting of four prospective, population-based birth cohorts that enrolled birthing mother and newborn pairs in Canada. Details of these cohorts have been described elsewhere (34). The current investigation focused on i). European-origin offspring from the population-based CHILD study who were selected for methylation analysis, ii). The Family Atherosclerosis Monitoring In early life (FAMILY) study that is predominately European-origin, and iii). The SouTh Asian biRth cohorT (START) study that is exclusively comprised of people who originated from the Indian subcontinent known as South Asians. The ethnicity of the parents was self-reported and recorded at baseline in all three cohorts. Biological samples, clinical assessments, and questionnaires were used to derive health phenotypes and an array of genetic, epigenetic, and metabolomic data. The superordinate goal of the NutriGen study is to understand how nutrition, environmental exposures, and physical health of mothers impact the health and early development of their offspring using a multi-omics approach.

### Methylation data processing and quality controls

Newborn cord blood samples were processed using two methylation array technologies. About half of the START samples and selected samples from CHILD were hybridized to the Illumina Human-Methylation450K BeadChip (HM450K) array, which covers CpGs in the entire genome (35) The raw methylation data were generated by the Illumina iScan software and separately pre-processed for START and CHILD using the “*sesame*” R package following pipelines designed for HM450K BeadChip (36). The FAMILY samples were profiled using a targeted array based on the Infinium Methylation EPIC designed by the Genetic and Molecular Epidemiology Laboratory (GMEL; Hamilton, Canada). The GMEL customized array includes ∼3000 CpG sites that were previously reported to associate with complex traits or exposures and was designed to maximize discovery while keeping the costs of profiling epigenome-wide DNA methylation down. The targeted methylation data were pre-processed using a customized quality control pipeline and functions from the “*sesame*” R package recommended for EPIC.

Pre-processed data were then used to derive the β-value matrix, where each column gives the methylation level at a CpG site as a ratio of the probe intensity to the overall probe intensity. Additional quality control filters were applied to the final beta-value matrices to remove samples with > 10% missing probes and CpG probes with >10% samples missing. Cross-reactive probes and SNP probes were removed as recommended for HM450 (37) and EPIC arrays (38,39). For CpG probes with a missing rate <10%, mean imputation was used to fill in the missing values. We further excluded samples that were either mismatches between reported sex and methylation-inferred sex or were duplicates. Finally, considering the low prevalence of smokers, we sought to reduce spurious associations by removing non-informative probes that were either all hypomethylated (β-value < 0.1) or hypermethylated (β-value > 0.9), which have been shown to have less optimal performance (40). A summary of the sample and probe inclusion/exclusion is shown in Supplementary Table 1. A detailed description of pre-processing and quality control steps is included in Supplementary Material.

Cell-type proportions (CD8T, CD4T, Natural Killer cells, B cells, monocytes, granulocytes, and nucleated red blood cells) were estimated following a reference-based approach developed for cord blood (41) and using R packages “*FlowSorted.CordBloodCombined.450k*” and “*FlowSorted.Blood.EPIC*”. All data processing and subsequent analyses were conducted in R v.4.1.0 (42).

### Phenotype data processing and quality controls

At the time of enrollment, all pregnant women completed a comprehensive questionnaire that collected information on prenatal diet, smoking, education, socioeconomic factors, physical activities and health as detailed previously (43,44). Maternal smoking history (0=never smoked, 1=quit before this pregnancy, 2=quit during this pregnancy, or 3=current smoker) was assessed during the second trimester (at baseline). Smoke exposure was measured as “number of hours exposed per week”. GDM was determined based on a combination of oral glucose tolerance test (OGTT), self-report, and reported diabetic treatments (insulin, pills, or restricted diet). For South Asian mothers in START, the same OGTT threshold as Born in Bradford (27,32) was used, while the International Association of the Diabetes and Pregnancy Study Groups (IASDPSG) criteria (45) for OGTT were used in CHILD and FAMILY cohorts. Mode of delivery (emergency c-section vs. other) was collected at the time of delivery.

Newborn length and weight were collected immediately after birth and extracted from medical chart. The newborns were then followed up at 1, 2, 3, and 5 years of age and provided basic anthropometric measurements, including height, weight, hip and waist circumference, BMI, sum of the skinfolds (triceps skinfold and subscapular skinfold). Additional phenotypes included smoking exposures (hours per week) at home, potential allergy based on mother reporting any of: eczema, hay fever, wheeze, asthma, food allergy (egg, cow milk, soy, other) for her child in FAMILY and START, and asthma based on mother’s opinion in CHILD (“In your opinion, does the child have any of the following? Asthma”).

### Phenotype and Methylation Data Consolidation

The current investigation examines the impact of maternal smoking or smoke exposure on DNA methylation derived from newborn cord blood in START and the two predominately European cohorts (CHILD and FAMILY). To maximize sample size in FAMILY and CHILD, we retained either self-identified or genetically confirmed Europeans based on available genetic data (Supplementary Table 1). The cohorts consist of representative population samples without enrichment for any clinical conditions, though only singleton mothers were invited to participate. For continuous phenotypes, an analysis of variance (ANOVA) using the *F*-statistics or a two-sample *t*-test was used to compare the mean difference across the three cohorts or two groups, respectively. For categorical phenotypes, a chi-square test of independence was used to compare the difference in frequencies of observed categories. Note that three of the categories under smoking history in the START cohort had expected cell counts less than 5, and was thus excluded from the comparison, the reported *p*-value was for CHILD and FAMILY.

The final analytical datasets, after combining the quality-controlled methylation data and phenotypic data, included 352, 411, and 504 mother-newborn pairs from CHILD, FAMILY, and START, respectively. Demographic characteristics and relevant covariates of the epigenetic subsample and the overall sample are summarized in Table 1 and Supplementary Table 2, respectively.

**Table 1.**
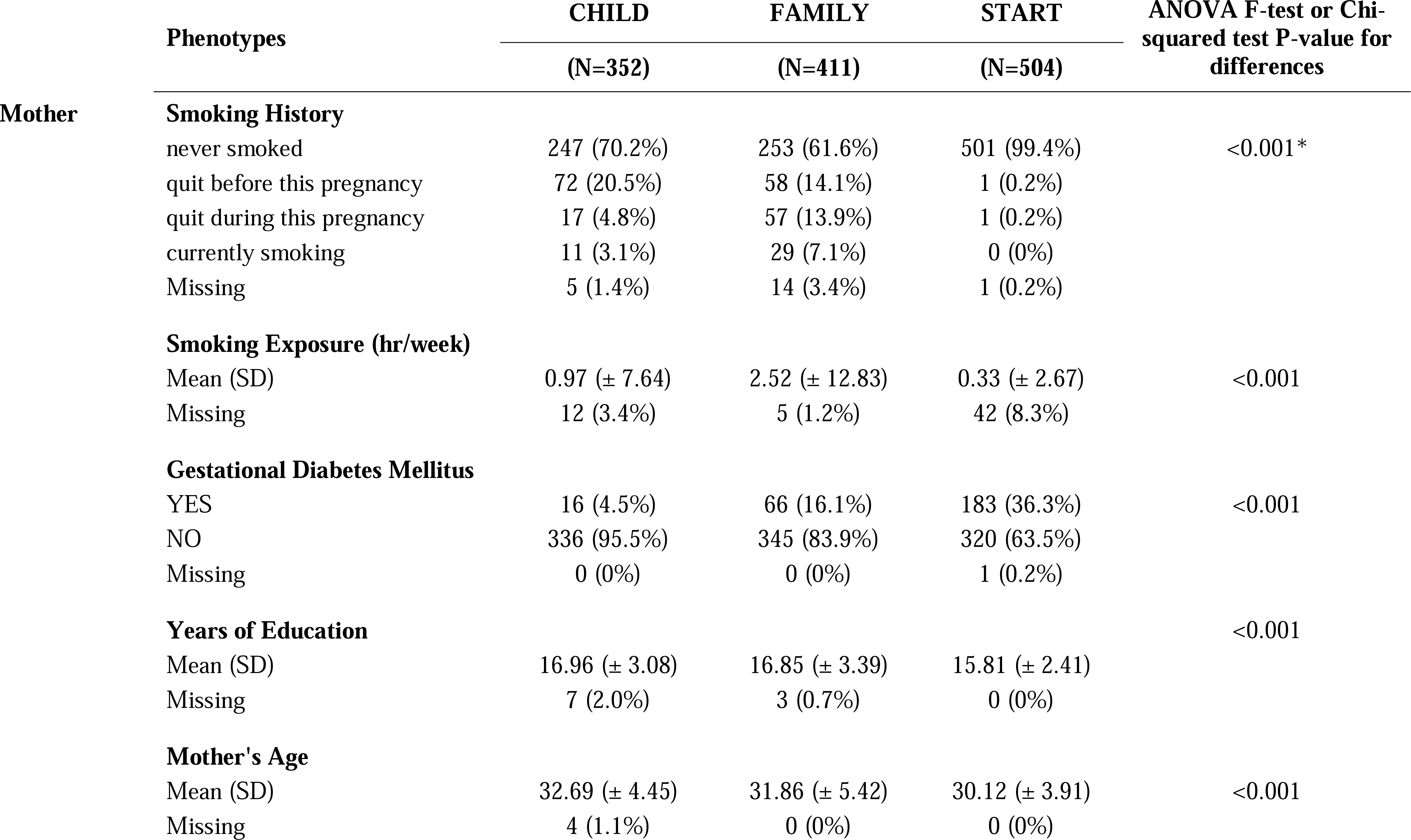
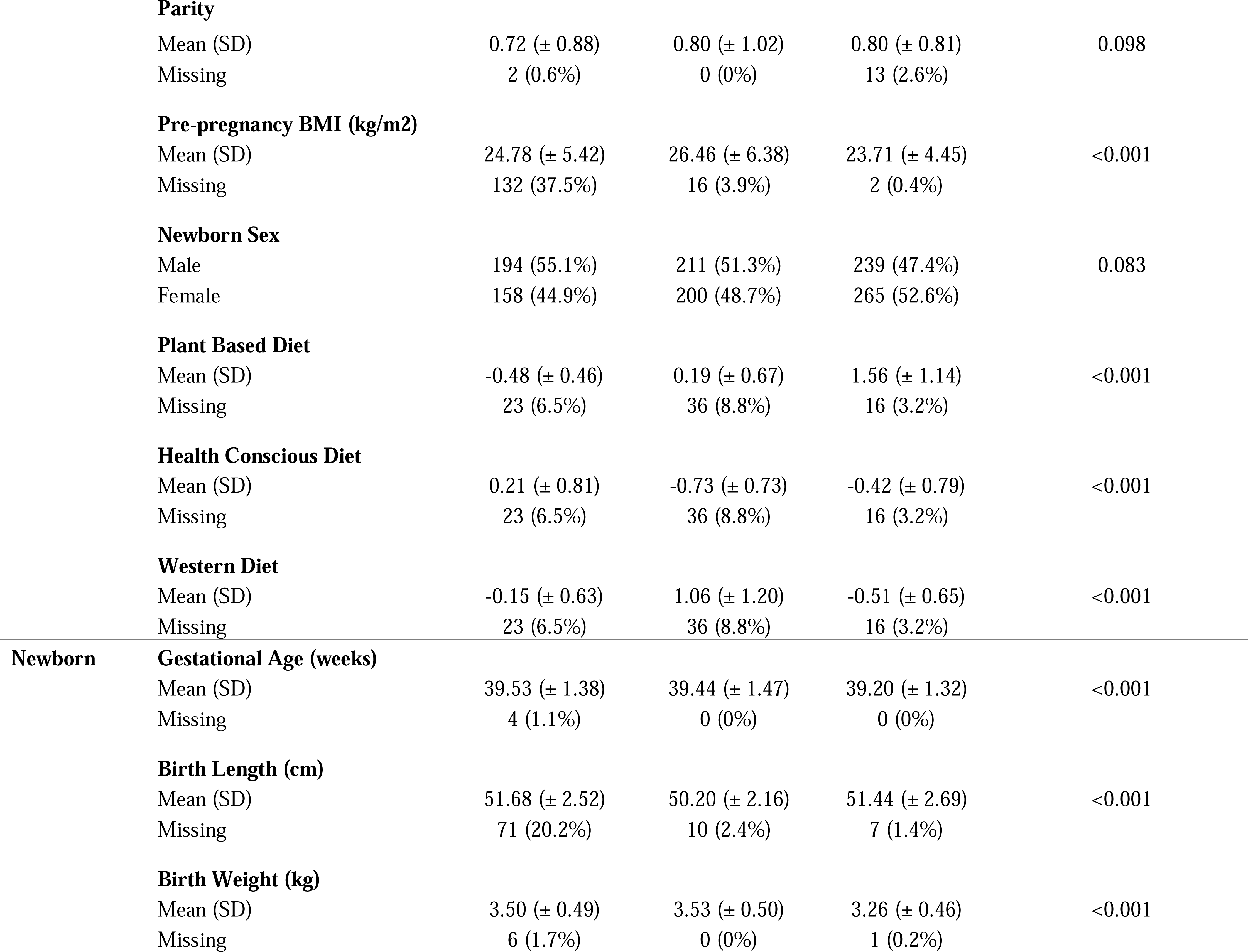
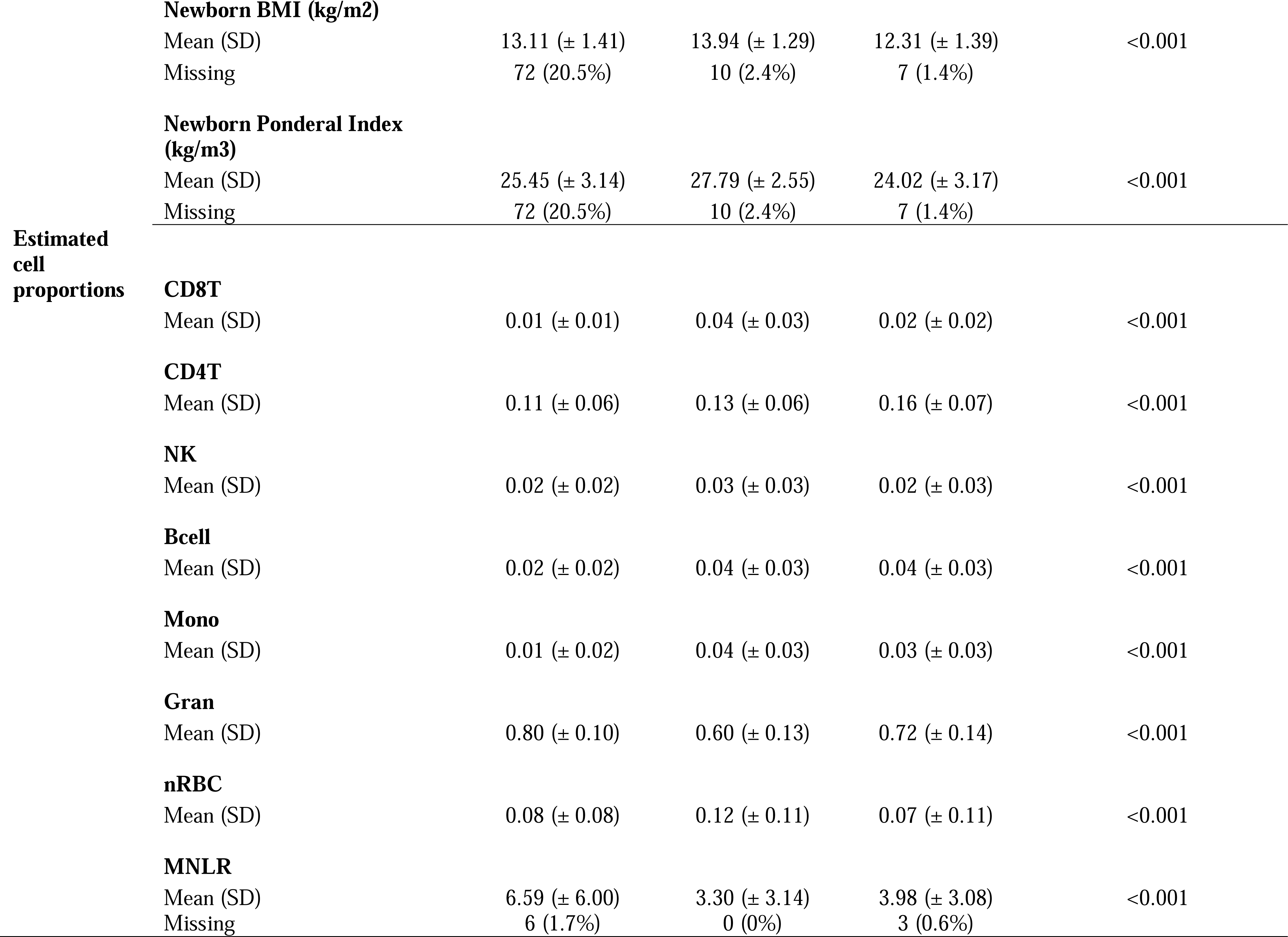
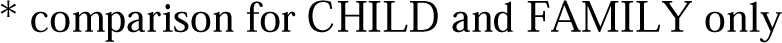
Characteristics of the epigenetic subsample (1,267 mother–newborn pairs) from the CHILD, FAMILY, START.

### Epigenome-Wide Association of Maternal Smoking in European Cohorts

Since there were no current smokers in START (Table 1), we tested the association between maternal smoking and differential methylated sites in FAMILY (# of CpG = 2,544) and CHILD (# of CpG = 200,050). The primary outcome variable was “current smoker”, defined by mothers self-identified as currently smoking during the pregnancy vs. those who never smoked or quit either before or during pregnancy. We also included a secondary outcome variable “ever smoker”, defined by mothers who are current smokers or have quit smoking vs. those who never smoked. A tertiary outcome was smoking exposure, measured by the number of hours per week reported by the expectant mothers, and was available in all cohorts. We summarized the type of analyses for different outcomes in Supplementary Table 3.

We first conducted a separate epigenetic association study in each cohort, testing the association between methylation β-values at individual CpGs and the smoking phenotype using either a logistic regression model for smoking status or a linear regression for smoking exposure as the outcome. The model adjusted for additional covariates including the estimated cord blood cell proportions, maternal age, social disadvantage index, which is a continuous composite measure of social and economic exposures (46), mother’s years of education, GDM, and parity. The smoking exposure variable was skewed, and a rank-based transformation was applied to mimic a standard normal distribution.

We then meta-analyzed association results for maternal smoking status in the European cohorts using an inverse variance-weighted fixed-effect model. The meta-analysis was conducted for 2,112 CpGs that were available in both CHILD (profiled using HM450K) and FAMILY (profiled using the targeted array). For the tertiary outcome, we conducted an inverse variance meta-analysis including START using both a fixed-effect model. For each EWAS or meta-analysis, the false discovery rate (FDR) adjustment was used to control multiple testing and we considered CpGs that passed an FDR-adjusted *p*-value < 0.05 to be relevant for maternal smoking.

### Using DNA Methylation to Construct Predictive Models for Maternal Smoking

We sought to construct a predictive model in the form of a methylation risk score (MRS) using reported associations of maternal smoking. The proposed solution adapted the existing lassosum method (47) that was originally designed for polygenic risk scores, where the matrix of SNP genotypes (*X*) can be conveniently replaced by the β-value matrix. For more details, see the Supplementary Material. Briefly, an objective function under elastic-net constraint was minimized to obtain the elastic-net solution y, where only summary statistics (b) and a scalar of the covariance between the β-values of the CpGs (X^I^X) are needed. The tuning parameters A and A_2_ were chosen by validating on the observed smoking history (as a continuous outcome) in CHILD that produced the most significant model. The optimized A_1_ and A_2_ were then used to create a final model that entails a list of CpGs and their corresponding weights, which were then used to calculate a MRS for maternal smoking in the FAMILY and START samples.

The summary statistics of the discovery EWAS were obtained from EWAS catalog (http://www.ewascatalog.org/) reported under “PubMed ID 27040690” by Joubert and colleagues (19). The summary statistics were restricted to the analysis “sustained maternal smoking in pregnancy effect on newborns adjusted for cell composition”. Of the 2620 maternal smoking CpGs that passed the initial screening, 2,107 were available in CHILD but only 128 were common to CHILD, FAMILY, and START. To evaluate whether the targeted GMEL-EPIC array design has comparable performance as the epigenome-wide array to evaluate the epigenetic signature of maternal smoking, a total of three MRSs were constructed, two using the 128 CpGs available in all cohorts – across the HM450K and targeted GMEL-EPIC arrays – and with either CHILD (n = 347 with non-missing smoking history) or FAMILY (n = 397 with non-missing smoking history) as the validation cohort, and another using 2,107 CpGs that were only available in CHILD and START samples with CHILD as the validation cohort. The validation model considered the continuous smoking history without modification as the outcome, while accounting for covariates, which included the estimated cord blood cell proportions, maternal age, social disadvantage index, mother’s years of education, GDM, and parity. Henceforth, we referred to these derived maternal smoking scores as the FAMILY targeted MRS, CHILD targeted MRS, and the HM450K MRS, respectively. To benchmark and compare with existing maternal smoking MRSs, we calculated the Reese score using 28 CpGs (48,49), Richmond score using 568 CpGs (49), Rauschert score using 204 CpGs (50), Joubert score using all 2,620 CpGs with evidence of association for maternal smoking (19), and finally a three-CpG score for air pollution (51). The details of these scores and score weight can be found in Supplementary Table 4.

### Statistical analysis

For each cohort, we contrasted the three versions of the derived scores using an analysis of variance analysis (ANOVA) along with pairwise comparisons using a two-sample *t*-test to examine how much information might be lost due to the exclusion of more than 10-fold CpGs at the validation stage, in all samples and in non-smokers. We also examined the correlation structure between all derived and external MRSs using a heatmap summarizing their pairwise Pearson’s correlation coefficient. Then, we compared the mean difference of each MRS score among smoking history using an ANOVA *F*-test and two-sample *t*-test to understand whether there was a dosage dependence in the cord blood DNAm signature of maternal smoking. Additionally, each score was tested against a binary outcome for current smoker vs. not, and two continuous measures for smoking history and weekly smoking exposure. The binary outcome was tested using a logistic regression model and the predictive performance was assessed using area under the receiver operating characteristic curve (AUC). The reported 95% confidence interval for each estimated AUC was derived using 2,000 bootstrap samples. The continuous outcome was examined using a linear regression model and its performance was quantified using the adjusted R^2^.

For the derived MRS, we empirically assessed whether a systematic difference existed in the resulting score with respect to all other derived scores. This was examined via pairwise mean differences between the HM450 and other scores using a two-sample *t*-test and an overall test of mean difference using an ANOVA *F*-test, among all samples and the subset of never smokers. Finally, we tested the association between each maternal smoking MRS and smoking phenotypes in mothers, as well as offspring phenotypes using a linear regression model, when applicable, adjusting for the child’s age at each visit. The association results were meta-analyzed for phenotypes with homogeneous effects across the cohorts using a fixed-effect model. An FDR adjustment was used to control the multiple testing of meta-analyzed associations between MRS and 25 (or 23, depending on the number of phenotypes available in the cohort) outcomes, and we considered association that passed an FDR-adjusted *p*-value < 0.05 to be relevant.

## Results

### Cohort Sample Characteristics

The analyses included 763 European mother-child pairs with cord blood DNAm data from the CHILD study (CHILD; *n* = 352)(52) and The Family Atherosclerosis Monitoring In earLY life (FAMILY; *n* = 411) study (43), and 503 South Asian mother-child pairs from The SouTh Asian biRth cohorT (START) study (44). We observed lower past smoking and missingness on smoking history among pregnant women in START as compared to CHILD or FAMILY using the epigenetic subsample (Table 1) and the overall sample (Supplementary Table 2). Pregnant women in START were significantly different from CHILD or FAMILY in that they were on average younger at delivery, had a lower BMI, and a higher rate of GDM, in line with other cohort studies in South Asian populations (53,54). As compared to START, newborn infants from CHILD and FAMILY had a longer gestational period, a higher birth weight, and a higher BMI at birth (Table 1; Supplementary Table 2). We observed no difference between cohorts in terms of parity or newborn sex in the epigenetic subsample (Table 1).

Within the European epigenetic subsample, of the 744 mother–newborn pairs with complete smoking history data, 40 (5.3%) newborns were exposed to current maternal smoking, which is on the lower end of the spectrum for the prevalence of smoking during pregnancy (9.2%-32.5%) among Canadians (55). In addition, mothers who smoked during pregnancy were on average younger, had fewer years of education, and had higher household exposure to smoking (Supplementary Table 6). However, there was no statistically significant difference between newborns exposed to current and none or previous smoking in terms of birth weight, birth length, gestational age, or estimated cord blood cell proportions.

### Epigenetic Association of Maternal Smoking in White Europeans

The two predominantly White European cohorts, FAMILY (*n* = 397) and CHILD (*n* = 347), contributed to the meta-analysis of maternal smoking for both the primary outcome of current smoking (Figure 2-A) and the secondary outcome of ever smoking (Supplementary Figure 1). The top associated CpGs with current maternal smoking were mapped to the growth factor independent 1 (*GFI1*) gene on chromosome 1, with cg12876356 as the lead (meta-analyzed effect = −1.11±0.22; meta-analyzed *p* = 2.6×10^-6^; FDR adjusted *p* = 0.006; Table 2). There were no CpGs associated with the ever-smoker status at an FDR of 0.05, though the top signal (cg09935388) was also mapped to the *GFI1* gene (Pearson’s r^2^ correlation with cg12876356 = 0.75 and 0.68 in CHILD and FAMILY, respectively; Supplementary Figure 1). The top associated CpG from the meta-analysis of smoking exposure (hours per week) in the European-origin cohorts (Figure 2-B) was cpg01798813 on chromosome 17, which was also associated with maternal smoking and was consistent in the direction of association (meta-analyzed effect = −0.18±0.04; meta-analyzed *p* = 1.4×10^-5^; FDR adjusted *p* = 0.04; Table 2). There was no noticeable inflation of empirical type I error in the association *p*-values from the meta-analysis, with the median of the observed association test statistic roughly equal to the expected median (Supplementary Figure 2). As a sensitivity analysis, we repeated the analysis for the continuous smoking exposure under rank transformation vs. raw phenotype for the associated CpG in *GFI1* and examined the regression diagnostics (Supplementary Material), and found that the model under rank-transformation deviated less from assumptions. Further, we observed consistency in the direction of association for the 128 CpGs that overlapped between our meta-analysis and the 2,620 CpGs with evidence of association for maternal smoking (19) (Supplementary Figure 3). Specifically, the Pearson’s correlation coefficient for maternal smoking and weekly smoking exposure was 0.72 and 0.60, respectively. The maternal smoking and smoking exposure EWASs in CHILD alone did not yield any CpGs after FDR correction (Supplementary Figure 4).

**Figure 1.**
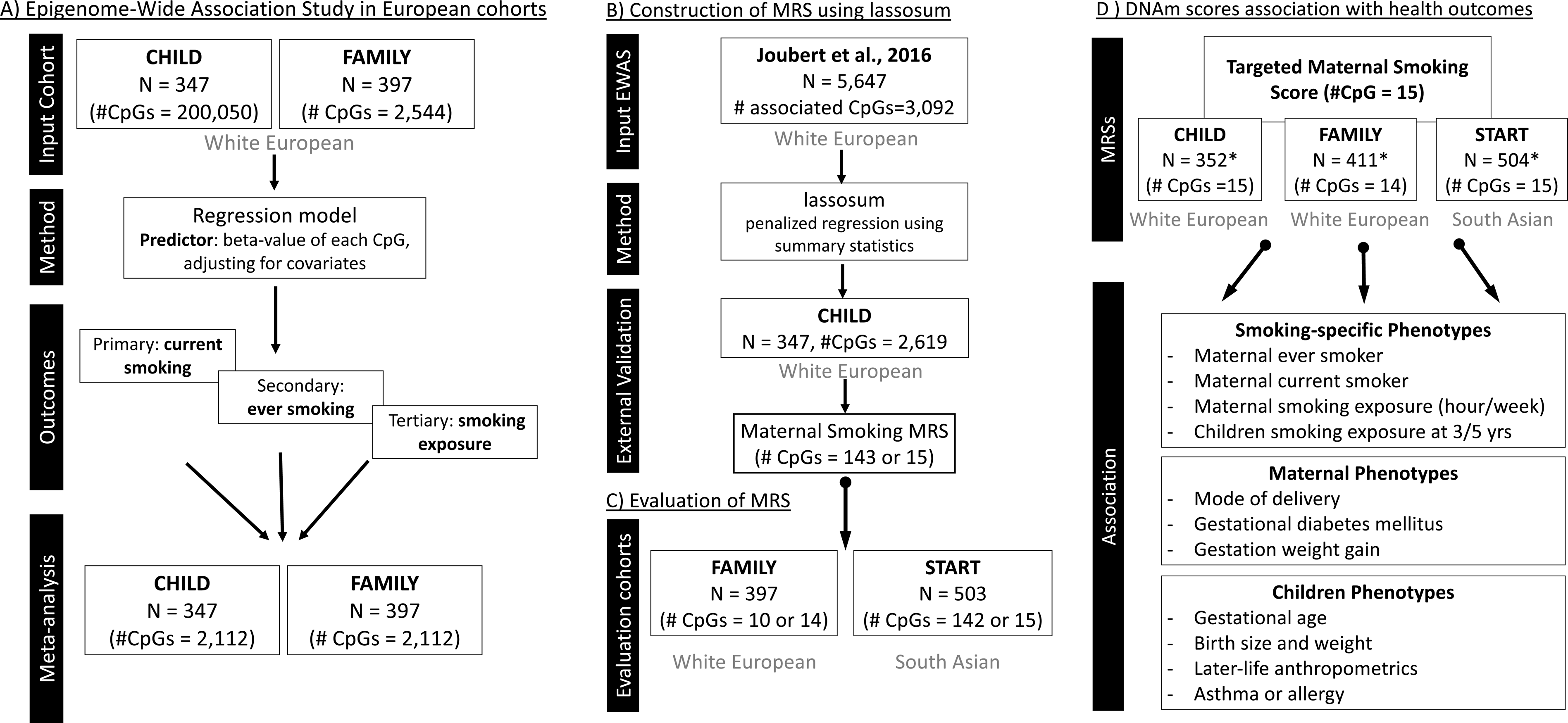
Schematic overview of the analytical pipeline for the cord blood DNAm maternal smoking score and association study. Figure 1-A) shows the epigenome-wide association studies conducted in the European cohorts (CHILD and FAMILY); Figure 1-B) illustrated the workflow for methylation risk score (MRS) construction using an external EWAS (Joubert et al., 2016) as the discovery sample and CHILD study as the external validation study, while Figure 1-C) demonstrates the evaluation of the MRS in two independent cohorts of white European (i.e. FAMILY) and South Asian (i.e. START). The validated MRS was then tested for association with smoking specific, maternal, and children phenotypes in CHILD, FAMILY, and START, as shown in Figure 1-D). * indicates cohort sample size including those with missing smoking history.

**Figure 2.**
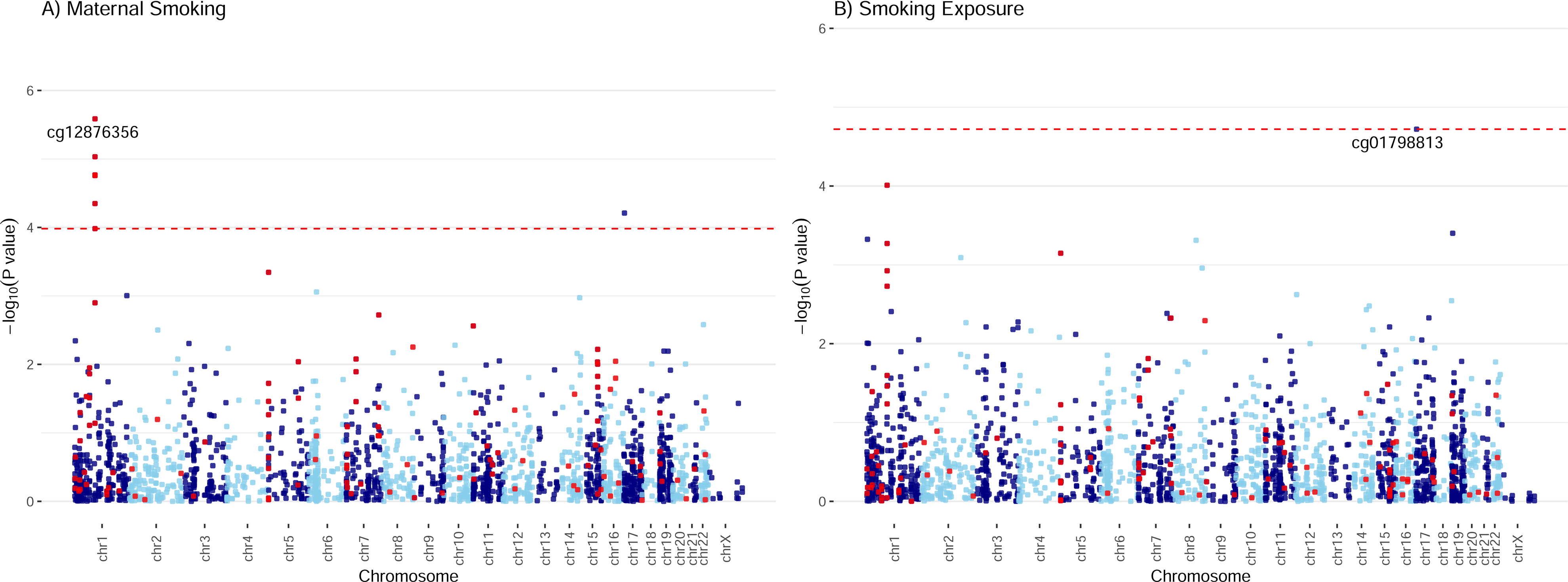
Manhattan plots of the meta-analyzed association between cord blood DNAm and maternal smoking in Europeans. Manhattan plots summarized the meta-analyzed association *p*-values between cord blood DNA methylation levels and current maternal smoking (A) or smoking exposure (B) at a common set of 2,114 CpG sites. The red line denotes the smallest -log10(*p*-value) that is below the FDR correction threshold of 0.05. The red dots represent established associations with maternal smoking reported in Joubert and colleagues (19).

**Table 2.**
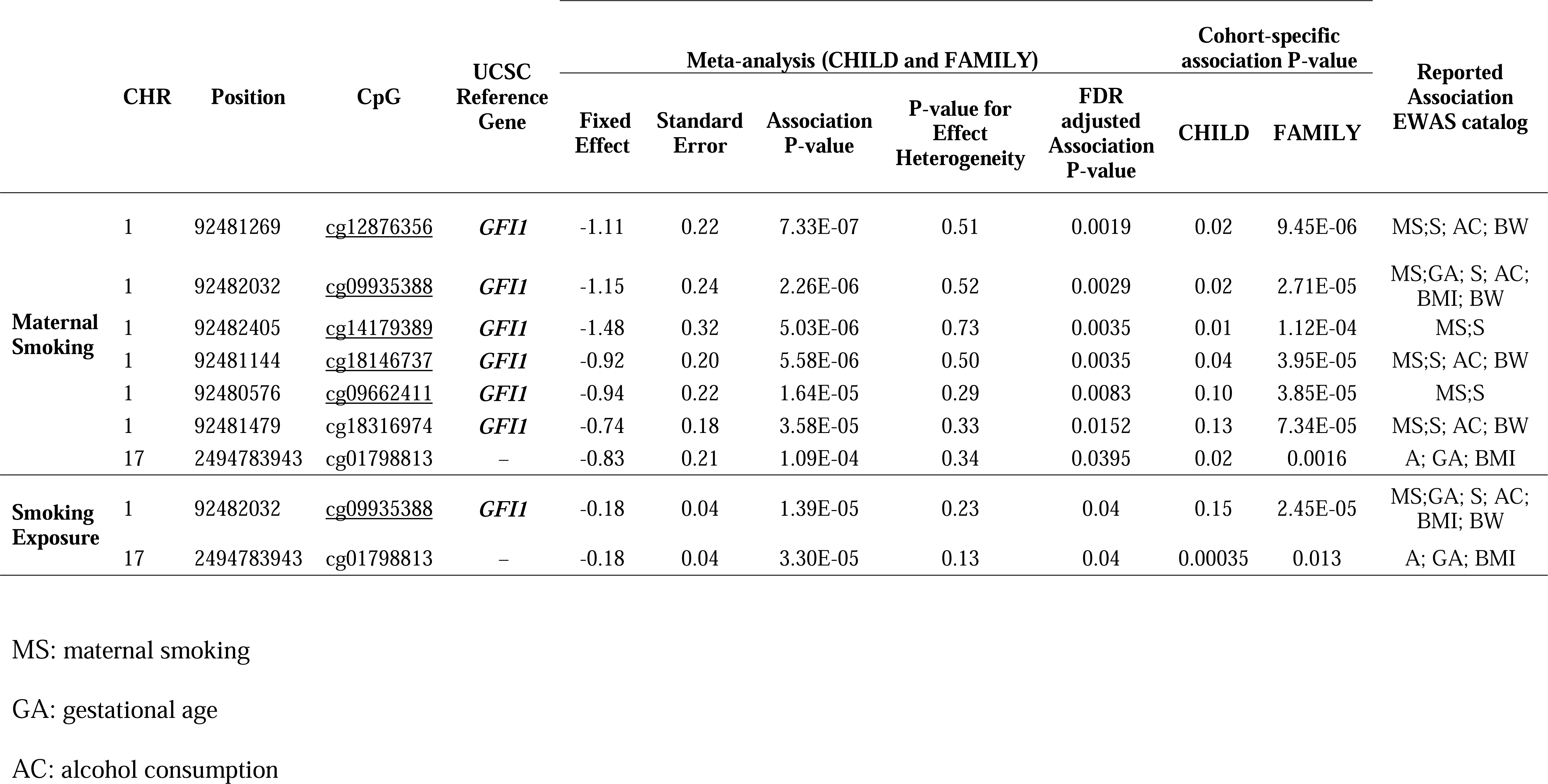
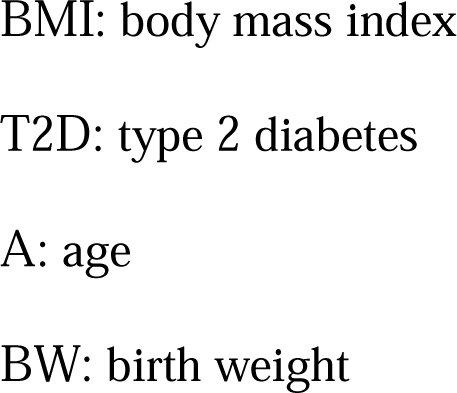
Meta-analysis results of association between CpGs and maternal smoking and smoking exposure that passed a marginal *p* < 0.05 threshold after the false discovery rate correction in European cohorts.

### Methylation Risk Score (MRS) Captures Maternal Smoking and Smoking Exposure

The final MRSs, validated using CHILD European samples (n = 347), included 15 and 143 CpG markers (Supplementary Table 7) from the targeted array and the epigenome-wide HM450 array, respectively. Both produced methylation scores that were significantly associated with maternal smoking history (ANOVA F-test *p*-values =1.0×10^-6^ and 2.4×10^-14^ in CHILD and 3.6×10^-16^ and <2.2×10^-16^ in FAMILY; Figure 3, Supplementary Figure 5), and the best among alternative scores for CHILD and FAMILY (Supplementary Table 5). With the exception of the air pollution MRS, which only contained 3 CpGs (Supplementary Table 5), all remaining scores were marginally associated with smoking history in both CHILD and FAMILY (Supplementary Figure 5) and correlated with each other (Supplementary Figure 6). In particular, scores that were derived using the Joubert EWAS as the discovery sample, including ours, had higher pairwise correlation coefficients across the birth cohorts, with many of the CpGs mapping to the same genes, such as *AHRR*, *MYO1G*, *GFI1*, *CYP1A1*, and *RUNX3*. There was no statistically significant difference in mean between the two scores in any of the three cohorts (two-sample t-test *p*s > 0.6) or among non-smokers (two-sample t-test *p*s> 0.6; Supplementary Figure 7). Since the HM450 score provides statistically more significant results in both CHILD and FAMILY with smoking history, despite the reduction in CpGs included (only 26 out of 143 CpGs present in FAMILY; Supplementary Table 5), we proceeded with the HM450 MRS model constructed using the 143 CpGs in subsequent analyses.

**Figure 3.**
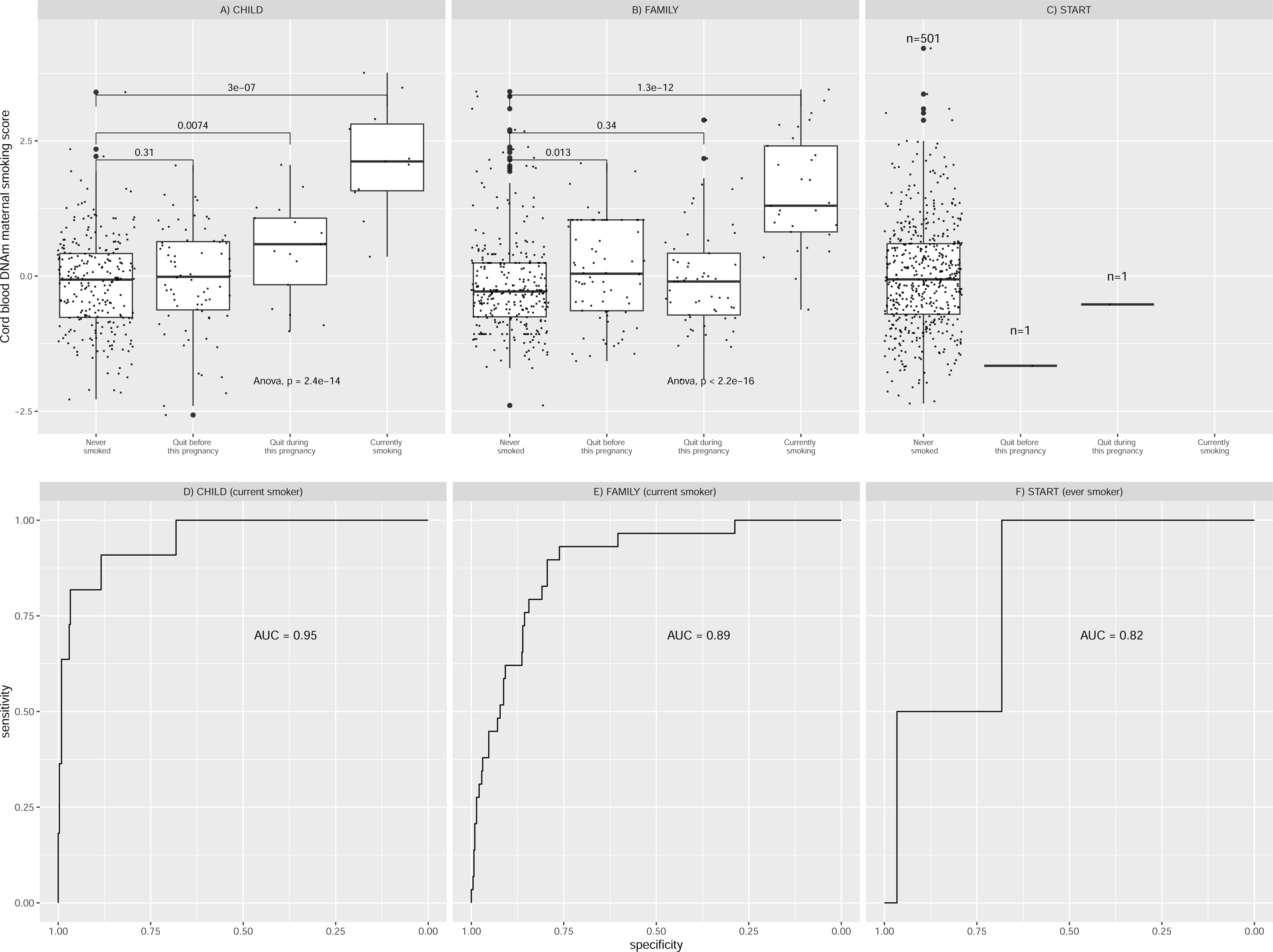
Relationships between maternal smoking MRS and maternal smoking history categories for each of the studies. Maternal smoking methylation score (y-axis) was shown as a function of maternal smoking history (x-axis) in levels of severity for prenatal exposure for CHILD (A), FAMILY (B), and START (C). Each severity level was compared to the never-smoking group and the corresponding two-sample *t*-test *p*-value was reported. The analysis of variance via an F-test *p*-value was used to indicate whether a mean difference in methylation score was present among all smoking history categories. The sample size for START cohort was provided due to the low counts in categories of any smoking. The area under the receiver operating characteristic curve (AUC) for each study was shown in the lower panel.

The HM450 MRS was significantly associated with maternal smoking history in CHILD and FAMILY (n = 397), but we failed to meaningfully validate the association in START (n = 503; Figure 3) – not surprisingly – due to the low number of ever-smokers (*n* = 2). A weak dose-dependent relationship between the MRS and the four categories of maternal smoking status in the severity of exposure ([0] = never smoked; [1] = quit before this pregnancy; [2] = quit during this pregnancy; [3] = currently smoking) was present in CHILD but was not replicated in FAMILY (Figure 3). The AUC for detecting current smokers were 0.95 (95% confidence interval: 0.89–1) and 0.89 (95% CI: 0.83–0.94) in CHILD and FAMILY (Figure 3), respectively, while the AUCs for detecting ever-smokers were 0.61 (95% CI: 0.54–0.67), 0.60 (95% CI: [0.55,0.69]; Supplementary Table 5), and 0.82 (95% CI: [0.55,1]; Figure 3), respectively. As a result, the epigenetic maternal smoking score was strongly associated with smoking status during pregnancy (OR=1.09, 95% CI: [1.07,1.10], *p*=1.96×10^-32^) in the combined European cohorts. Meanwhile, the maternal smoking MRS was significantly associated with increased number of hours exposed to smoking per week in the two White European cohorts (1.93±0.33 hours per 1 unit of increase in MRS, FDR adjusted *p* = 1.2×10^-7^; Supplementary Table 8; cohort specific *p*=5.4×10^-5^ in CHILD and *p*=2.3×10^-5^ in FAMILY; Table 3), but not in the South Asian birth cohort (*p* = 0.58; Table 3).

**Table 3.**
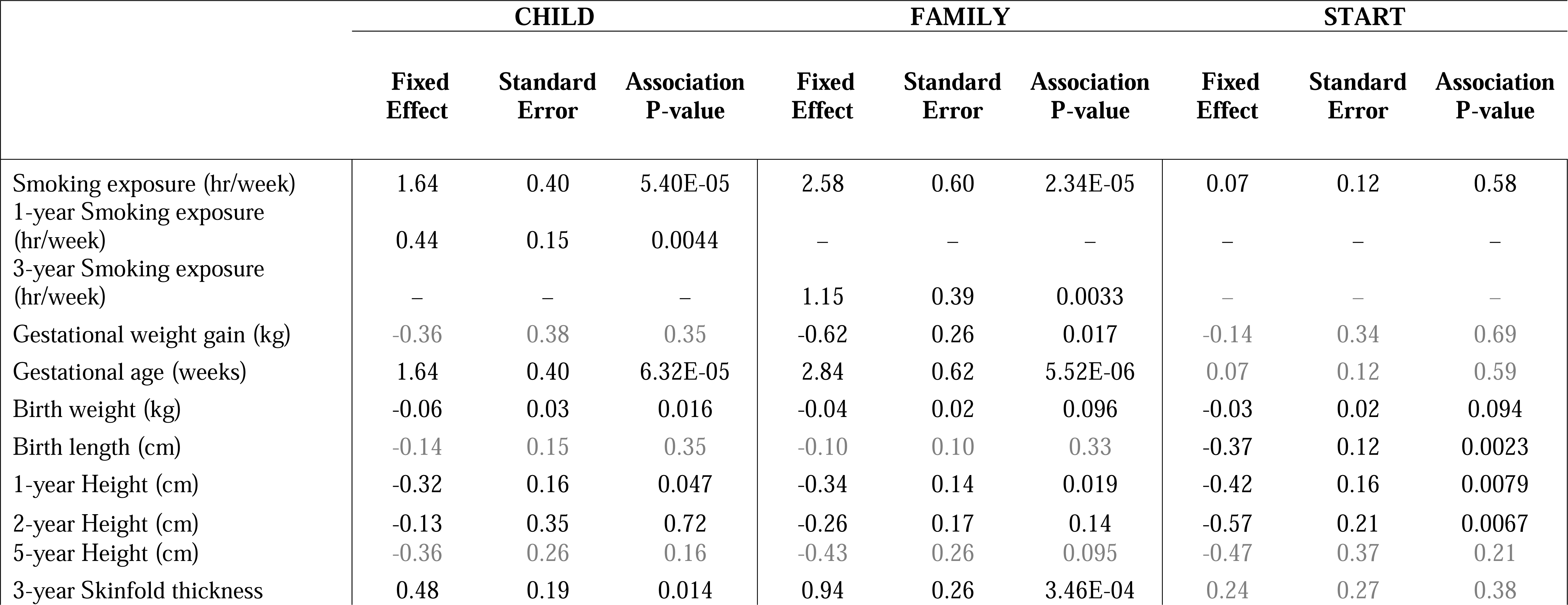
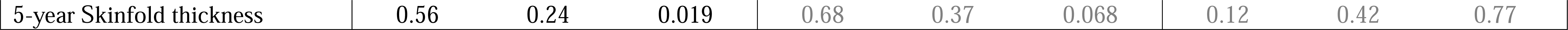
Significant associations between maternal smoking methylation risk score and phenotypes in CHILD, FAMILY and START.

|  | CHILD |  |  | FAMILY |  |  | START |  |  |
| --- | --- | --- | --- | --- | --- | --- | --- | --- | --- |
|  | Fixed Effect | Standard Error | Association P-value | Fixed Effect | Standard Error | Association P-value | Fixed Effect | Standard Error | Association P-value |
| Smoking exposure (hr/week) | 1.64 | 0.40 | 5.40E-05 | 2.58 | 0.60 | 2.34E-05 | 0.07 | 0.12 | 0.58 |
| 1-year Smoking exposure (hr/week) | 0.44 | 0.15 | 0.0044 | – | – | – | – | – | – |
| 3-year Smoking exposure (hr/week) | – | – | – | 1.15 | 0.39 | 0.0033 | – | – | – |
| Gestational weight gain (kg) | -0.36 | 0.38 | 0.35 | -0.62 | 0.26 | 0.017 | -0.14 | 0.34 | 0.69 |
| Gestational age (weeks) | 1.64 | 0.40 | 6.32E-05 | 2.84 | 0.62 | 5.52E-06 | 0.07 | 0.12 | 0.59 |
| Birth weight (kg) | -0.06 | 0.03 | 0.016 | -0.04 | 0.02 | 0.096 | -0.03 | 0.02 | 0.094 |
| Birth length (cm) | -0.14 | 0.15 | 0.35 | -0.10 | 0.10 | 0.33 | -0.37 | 0.12 | 0.0023 |
| 1-year Height (cm) | -0.32 | 0.16 | 0.047 | -0.34 | 0.14 | 0.019 | -0.42 | 0.16 | 0.0079 |
| 2-year Height (cm) | -0.13 | 0.35 | 0.72 | -0.26 | 0.17 | 0.14 | -0.57 | 0.21 | 0.0067 |
| 5-year Height (cm) | -0.36 | 0.26 | 0.16 | -0.43 | 0.26 | 0.095 | -0.47 | 0.37 | 0.21 |
| 3-year Skinfold thickness | 0.48 | 0.19 | 0.014 | 0.94 | 0.26 | 3.46E-04 | 0.24 | 0.27 | 0.38 |
| 5-year Skinfold thickness | 0.56 | 0.24 | 0.019 | 0.68 | 0.37 | 0.068 | 0.12 | 0.42 | 0.77 |

Among individuals who had never smoked, no statistically significant mean difference was observed in the distribution of the combined methylation score between South Asian and European cohorts (Supplementary Table 9). These results provided empirical support for the portability of an European-derived maternal smoking methylation score to South Asian populations.

### Association between MRS and other phenotypes

The maternal smoking MRS was consistently associated with increasing weekly smoking exposure in children reported by mothers at the 1-year visit (0.44±0.15, *p* = 0.0044; Table 3) in CHILD, and at 3-year visit (0.86±0.26, *p* = 0.0037; Table 3) in FAMILY, but not in START as all mothers reported non-exposure to smoking in children. A higher maternal smoking MRS was significantly associated with smaller birth size (−0.37±0.12, *p* = 0.0023; Table 3) and height at 1, 2, and 5 year visits in the South Asian cohort (Table 3). We observed similar associations with body size in the white European cohorts (heterogeneity *p*-values> 0.2), collectively, the MRS was associated with a smaller birth size (−0.22±0.07, *p*=0.0016; FDR adjusted *p* = 0.019; Supplementary Table 8) in the combined European and South Asian cohorts. Meanwhile, a higher maternal smoking MRS was also associated with a lower birth weight (−0.043±0.013, *p* = 0.001; FDR adjusted *p* = 0.011; Supplementary Table 8) in the combined sample, though the effect was weaker in START (−0.03±0.02; *p* = 0.094; Table 3) as compared to the white European cohorts.

The meta-analysis revealed no heterogeneity in the direction nor the effect size of associations for body size and weight between populations at birth or at later visits (heterogeneity *p*-values = 0.16–1; Supplementary Table 8). The association between the MRS and several children phenotypes, including height or length, weight, and skinfolds, appeared to persist with similar estimated effects throughout early developmental years (Supplementary Table 8), albeit the most significant effects were at birth, and the significance attenuated at later visits. We did not find any association with self-reported allergy or asthma in children at later visits (Supplementary Table 8). Further, there was no evidence of association between the MRS and any maternal outcomes (Supplementary Table 8).

## Discussion

We examined the epigenetic signature of maternal smoking and smoking exposure using newborn cord blood samples from predominately European-origin and South Asian cohorts via two strategies: an individual CpG-level EWAS approach, and a multivariate approach in the form of a methylation score. The EWAS results replicated the association between maternal smoking and CpGs in the *GFI1* gene that is well described in the literature with respect to smoking (15,17), maternal smoking (19,20,56,57), and birth weight (23). In the latter case, we observed a significant association with maternal smoking history and smoking exposure in European-origin newborns. Further, we noted a weak dose-dependent relationship between maternal smoking history and the methylation score in one European cohort (CHILD) but this was not replicated in the other (FAMILY). Since the timing and duration of maternal smoking during pregnancy were not directly available, these differences could play a role in the magnitude and specificity of DNA methylation changes in cord blood. Finally, the significant association of the MRS with the newborn health metrics in START, in the absence of mothers’ active smoking, could be the result of underreporting of smoking, poor recall of the time of quitting, and/or due to air pollution exposure (58), leading to oxidative stress. This suggests that our cord blood DNAm signature of maternal smoking is perhaps not unique to cigarette smoking, but captures similar biochemical responses, for example, via the aryl hydrocarbon receptor (59,60). Our observation that a higher MRS was associated with lower birth weight and smaller birth length in both ethnic populations is thus consistent with the established link between oxidative stress and metabolic syndrome (61).

Contrary to DNA methylation studies of smoking in adults, where whole blood is often used as a proxy tissue, there are multiple relevant tissues for maternal smoking during pregnancy, including the placenta of the mother, newborn cord blood, and children’s whole blood. However, methylation changes measured in whole blood or placenta of the mother, or cord blood of infants showed substantially different patterns of association signals (62). There are several advantages of using a cord blood based biomarker from the DoHaD perspective. Firstly, cord blood provides a direct reflection of the *in utero* environment and fetal exposure to maternal smoking. Additionally, since cord blood is collected at birth, it eliminates potential confounding factors such as postnatal exposures that may affect maternal blood samples. Furthermore, studying cord blood DNAm allows for the assessment of epigenetic changes specifically relevant to the newborn, offering valuable information on the potential long-term health implications. Meanwhile, methylation signals are known to be tissue-specific, thus it would be of interest for future research to combine differential methylation patterns from all relevant tissue to assess the immediate and long-term effects of maternal smoking. Another direction to further this line of research is to explore postnatal factors that mitigate prenatal exposures, for example, breastfeeding, which has been shown to have a protective effect against maternal tobacco smoking (63). Indeed, more research is necessary to understand the critical periods of exposure and the dose-response relationship between maternal smoking and cord blood DNA methylation changes. Ongoing efforts to monitor the offspring and collect data in the next decade are in progress to establish the long-term association between maternal smoking and cardio-metabolic health (43,44). As such, the constructed MRS can facilitate future research in child health and will be included as part of the generated data for others to access.

The strengths of this report include ethnic diversity, and fine phenotyping in a prospective and harmonized way with follow-up at multiple early childhood stages. This work is the first major multi-ancestry study that utilizes methylation scores to study maternal smoking and examines their portability from European-origin populations to South Asians. The use of MRS, as compared to individual CpGs, is a powerful tool to systematically investigate the influence of DNA methylation changes and whether it has lasting functional consequences on health outcomes. Our results converge with previous findings that epigenetic associations of maternal smoking are associated with newborn health, and add to the small body of evidence that these relationships extend to non-European populations and that different ancestral populations can experience the early developmental periods differently.

A few limitations should be mentioned. In the context of existing epigenetic studies of maternal smoking, we were not able to replicate signals in other well-reported genes such as *AHRR*, *CYP1A1*, and *MYO1G*, however, the MRS was able to pick up signals from these genes (Supplementary Table 7). This could be due to several reasons. First, the customized array with a limited number of CpGs (<3,000) was designed in 2016 and many large EWASs on smoking and maternal smoking conducted more recently had not been included. Nonetheless, we have shown that from a multivariate perspective, the MRS constructed using a targeted approach that was carefully designed can be equally powerful with the advantage of being cost-effective. Second, contrary to existing EWASs where the methylation values are typically treated as the outcome, and the exposure, such as smoking, as the predictor; we reversed the regression such that the methylation levels were the predictors and smoking exposure as the outcome. This reverse regression approach is robust and our choice to reverse the regression was motivated by the goal of constructing a smoking score that combines the additive effects at multiple CpGs, which would otherwise be unfeasible. Third, systematic ancestral differences in DNA methylation patterns had been shown to vary at individual CpGs in terms of their association with smoking (30). Converging with this conclusion, we also found the association with *GFI1* to be most consistent after adjusting for cell composition. Fourth, while it would be of interest to examine a broader range of health outcomes in children, such as lung health and allergies, we were unable to acquire and standardize this information across different cohorts. This aspect should be considered in future study designs. Finally, maternal smoking is often associated with other confounding factors, such as socioeconomic status, other lifestyle behaviours, and environmental exposures. While we have done our best to control for well-known confounders that were available by study design, as in all observational studies, we could not account for unknown confounding effects. Finally, in recent years, maternal smoking is on a decline as a result of changes in social norms and public health policies (64). This is also consistent with the lower smoking rates observed in our European cohorts (CHILD and FAMILY). Given the proportion of current smokers, the effective sample size for a direct comparison between CHILD and FAMILY, i.e. equivalently-powdered sample size of a balanced (50% cases, 50% controls) design, were 41.7 and 104.7, respectively. While CHILD had a lower effective sample size, we ultimately chose it for validating the methylation score to better cover the CpGs that were significant in the discovery EWAS. A larger validation study will likely further boost the performance of the methylation score and be considered in future research.

In conclusion, the epigenetic maternal smoking score we constructed was strongly associated with smoking status during pregnancy and self-reported smoking exposure in White Europeans, and with smaller birth size and lower birth weight in the combined South Asian and White European cohorts. The proposed cord blood epigenetic signature of maternal smoking has the potential to identify newborns who were exposed to maternal smoking *in utero* and to assess the long-term impact of smoking exposure on offspring health. In South Asian mothers with minimal smoking behaviour, the relationship between the methylation score and negative health outcomes in newborns is still apparent, indicating that DNA methylation response is sensitive to smoking exposure, even in the absence of active smoking.

## Supplementary Material

**Suppl. Table 1. Quality controls for the inclusion/exclusion of samples and methylation probes.**

**Suppl. Table 2. Characteristics of the overall sample include 5176 mother–newborn pairs from the CHILD, FAMILY, and START cohorts.**

**Suppl. Table 3. A summary of available analyses and outcome variables in each cohort.**

**Suppl. Table 4. A summary of the DNAm maternal smoking score derivation design and results.**

**Suppl. Table 5. Score weights for external DNAm maternal smoking scores.**

**Suppl. Table 6. Characteristics of the epigenetic subsample from CHILD and FAMILY cohorts stratified by smoking status.**

**Suppl. Table 7. A summary of CpGs that contribute to the DNAm maternal smoking scores and their weights.**

**Suppl. Table 8. Association between maternal smoking methylation risk score and phenotypes in CHILD, FAMILY and START.**

**Suppl. Table 9. Summary of mean difference in methylation risk scores between studies in overall samples and those never smoked.**

**Supplementary Figure 1. Manhattan plots of the meta-analyzed association between cord blood DNA methylation and ever maternal smoking in the combined European cohorts.**

The meta-analyzed association *p*-values for ever maternal smoking and methylation levels at 2,114 CpG sites were summarized in the Manhattan plot. Ever maternal smoking was defined to compare those who were currently smoking or quitted before or during this pregnancy vs. those never smoked. The red line denotes the smallest -log10(*p*-value) that is below the FDR correction threshold of 0.05. The red dots represent established associations with maternal smoking reported in Joubert and colleagues (19).

**Supplementary Figure 2. Quantile-quantile plots of the meta-analyzed association between cord blood DNA methylation and maternal smoking history, smoking exposure in the combined European cohorts.**

Quantile-quantile plots summarized the association *p*-values between cord blood DNA methylation levels and current maternal smoking (A) or ever maternal smoking (B) or weekly smoking exposure (C) at 2,114 CpG sites. The red line (y=x) is the line of reference and the genomic inflation factor, calculated as the ratio between the observed median and the theoretical median of the association test statistics, was annotated for each outcome.

**Supplementary Figure 3. Scatterplots of meta-analyzed association effects for maternal smoking history or smoking exposure and reported effects of maternal smoking.**

Panel A) is the scatterplot of meta-analyzed effects for maternal smoking in the combined CHILD and FAMILY cohorts (x-axis) vs. reported effects for maternal smoking in Joubert et al., 2016 (y-axis) for all CpGs present in CHILD, FAMILY, and Joubert et al., 2016 (# CpGs = 128); Panel B) is the scatterplot of meta-analyzed effects for weekly smoking exposure in the combined CHILD and FAMILY cohorts (x-axis) vs. reported effects for maternal smoking in Joubert et al., 2016 (y-axis) for all CpGs present in CHILD, FAMILY, and Joubert et al., 2016 (# CpGs = 128). The solid gray line is the best fitted line (95% confidence interval shown as the shaded area) for the linear relationship between the effect sizes and the dashed gray line represents the reference of y=x. (19)

**Supplementary Figure 4. Manhattan plots of the Epigenome-wide associations between cord blood DNAm and maternal smoking history, smoking exposure in CHILD.**

Manhattan plots summarized the association *p*-values between cord blood DNA methylation levels and current maternal smoking (A) or ever maternal smoking (B) or weekly smoking exposure (C) at 200,050 CpG sites. The red line denotes the smallest -log10(*p*-value) that is below the FDR correction threshold of 0.05. The red dots represent established associations with maternal smoking reported in Joubert and colleagues (19).

**Supplementary Figure 5. A comparison of results for derived and external maternal smoking MRSs.**

Maternal smoking methylation score (y-axis) was shown as a function of maternal smoking history (x-axis) in levels of severity ([0] = never smoked; [1] = quit before this pregnancy; [2] = quit during this pregnancy; [3] = currently smoking) for prenatal exposure for each study. **The scores shown were validated in 1) CHILD, 2) CHILD but restricted to CpGs that were also present on the targeted array, 3) FAMILY using CpGs on the targeted array.** Each severity level was compared to the never smoking group and the corresponding two sample *t*-test *p*-value was reported. An omnibus test *p*-value to test whether a mean difference in methylation score was present among all smoking history categories.

**Supplementary Figure 6. A heatmap of correlation between derived and external maternal smoking MRSs.**

This heatmap illustrates the pairwise correlation between MRSs calculated in A) CHILD, B) FAMILY, and C) START. Each cell represents the correlation coefficient, ranging from −1 to 1, indicating the strength and direction of the association. A value of 1 signifies a perfect positive correlation, while −1 indicates a perfect negative correlation. Values closer to 0 suggest no correlation. The color gradient from deep blue (strong negative correlation), through white (no correlation), to deep red (strong positive correlation), visually encodes the strength of these relationships. The scores in the black box were derived using lassosum and internally validated.

**Supplementary Figure 7. Comparison of all methylation scores stratified by study.**

The boxplots captured the standardized maternal smoking methylation scores (y-axis) stratified by study. The top panels summarized results for all samples in CHILD, FAMILY, and START, while the bottom panels summarized results for only those in CHILD, FAMILY, and START that never smoked. The *p*-values indicate the significance for a mean difference for each pairwise comparison between the HM450K score validated in CHILD with other scores using two-sample *t*-tests.

## Acknowledgements

We express our sincere gratitude to all the participating families and the START, FAMILY, and CHILD study teams, including interviewers, nurses, computer and laboratory technicians, clerical workers, research scientists, volunteers, managers, and receptionists.

We would like to acknowledge the Genetic and Molecular Epidemiology Laboratory (GMEL), an associate of Hamilton Health Sciences and McMaster University, for their indispensable contributions to this work. The technical staff of GMEL conducted all epigenetic profiling, including sample processing and other technical operations.

We thank the members of the Nutrigen Alliance for providing the data: Sonia S. Anand; Stephanie A. Atkinson; Meghan Azad; Allan B. Becker; Jeffrey Brook; Judah A Denburg; Dipika Desai; Russell J. de Souza; Milan K. Gupta; Michael Kobor; Diana L. Lefebvre; Wendy Lou; Piushkumar J. Mandhane; Sarah McDonald; Andrew Mente; David Meyre; Theo J. Moraes; Katherine M. Morrison; Guillaume Paré; Malcolm R. Sears; Padmaja Subbarao; Koon K. Teo; Stuart E. Turvey; Julie Wilson; Salim Yusuf; Gita Wahi; Michael A. Zulyniak.

This study was funded by the Canadian Institutes of Health Research Metabolomics Team Grant: MWG-146332. Dr. Anand is supported by a Tier 1 Canada Research Chair in Ethnicity and CVD and Heart, Stroke Foundation Chair in Population Health, a grant from the Canadian Partnership Against Cancer, Heart and Stroke Foundation of Canada and Canadian Institutes of Health Research. Dr. Azad is supported by a Tier 2 Canada Research Chair in the Developmental Origins of Chronic Disease.

## Data availability statement

The summary statistics used to construct methylation risk scores are available from EWAS catalog at http://www.ewascatalog.org/?trait=maternal%20smoking%20in%20pregnancy with additional filters of PubMID 27040690 and analysis on “Sustained maternal smoking in pregnancy effect on newborns adjusted for cell composition”.

Summary statistics generated in the current study, including a total of 8 primary association studies (three smoking phenotypes in three cohorts) and 3 sets of meta-analyzed results in Europeans are available upon request. All scripts to reproduce and validate the predictive model can be found at https://github.com/WeiAkaneDeng/EpigeneticResearch/tree/main/MaternalSmoking.

## Conflicts of interest

No conflict of interest.

## Ethics Statement

Ethical approval was obtained independently from the Hamilton Integrated Research Ethics Board: CHILD (REB 07–2929), FAMILY (REB 02–060), and START (REB 10–640). CHILD was additionally approved by the respective Human Research Ethics Boards at McMaster University, the Universities of Manitoba, Alberta, and British Columbia, and the Hospital for Sick Children. Legal guardians of each participant provided written informed consent. Written informed consent was obtained from the parent/guardian (participating mother) for each study separately. We also have now obtained additional ethics board approval from HiREB (REB 16592) for using the data from the three cohorts together without additional consent from the participants.

## Notes

### Competing Interest Statement

The authors have declared no competing interest.

### Author Declarations

Ethics approval: McMaster University. Ethical approval was obtained independently for all studies from the Hamilton Integrated Research Ethics Board CHILD (REB 07-2929), FAMILY (REB 02-060), START (REB 10-640).

### Summary of Updates

In the current revision, we have taken the recommendations by reviewers to 1) better illustrate the analytical flow and readability of figures; 2) reporting standards of p-values and other statistical metrics of associations; and 3) improved wording on the generalizability of maternal smoking score across cohorts due to low exposure in South Asians.

