## Supplementary figures and images for "Maternal smoking DNA methylation risk score associated with health outcomes in offspring of European and South Asian ancestry"

### Supplementary Figure 1

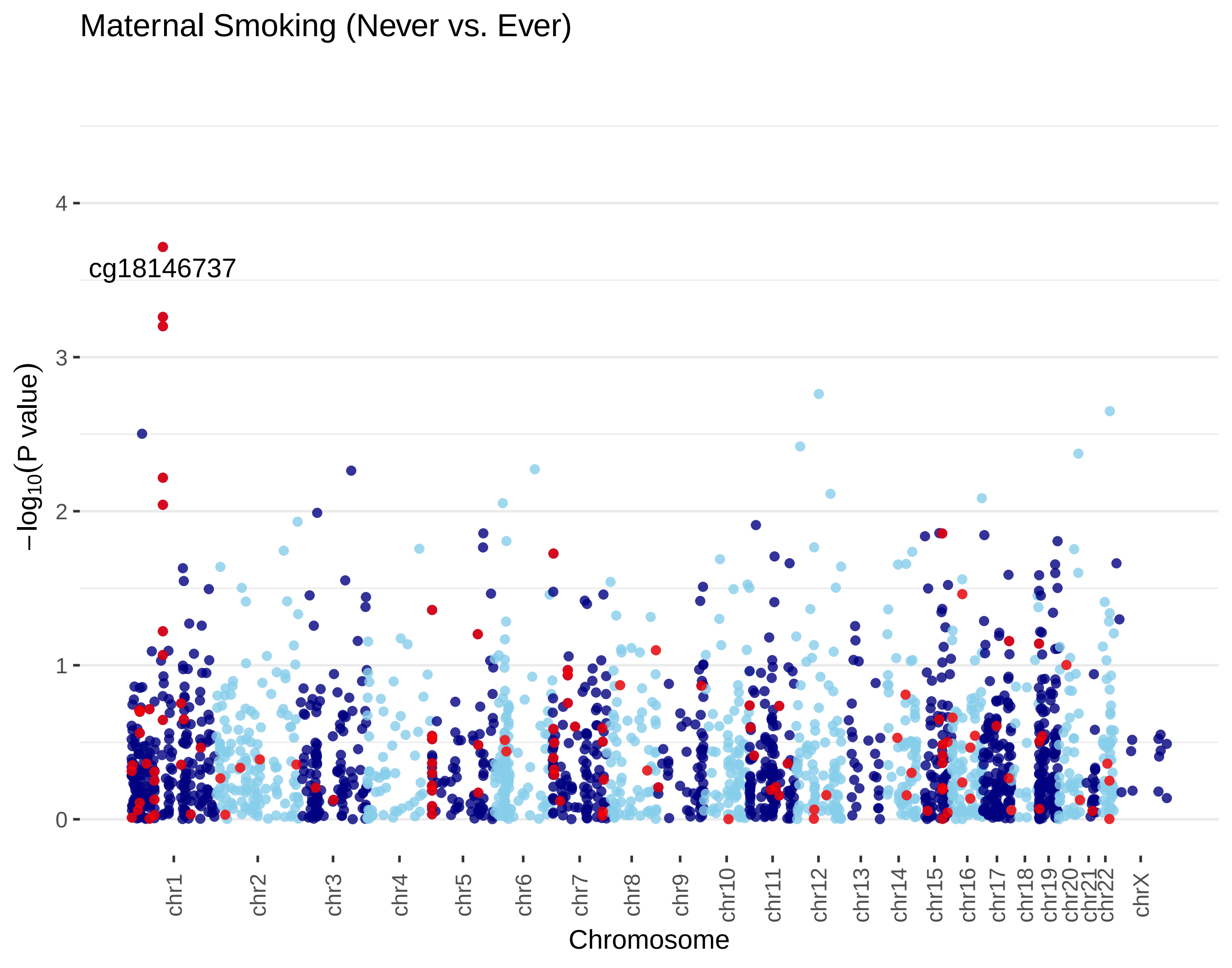

### Supplementary Figure 2

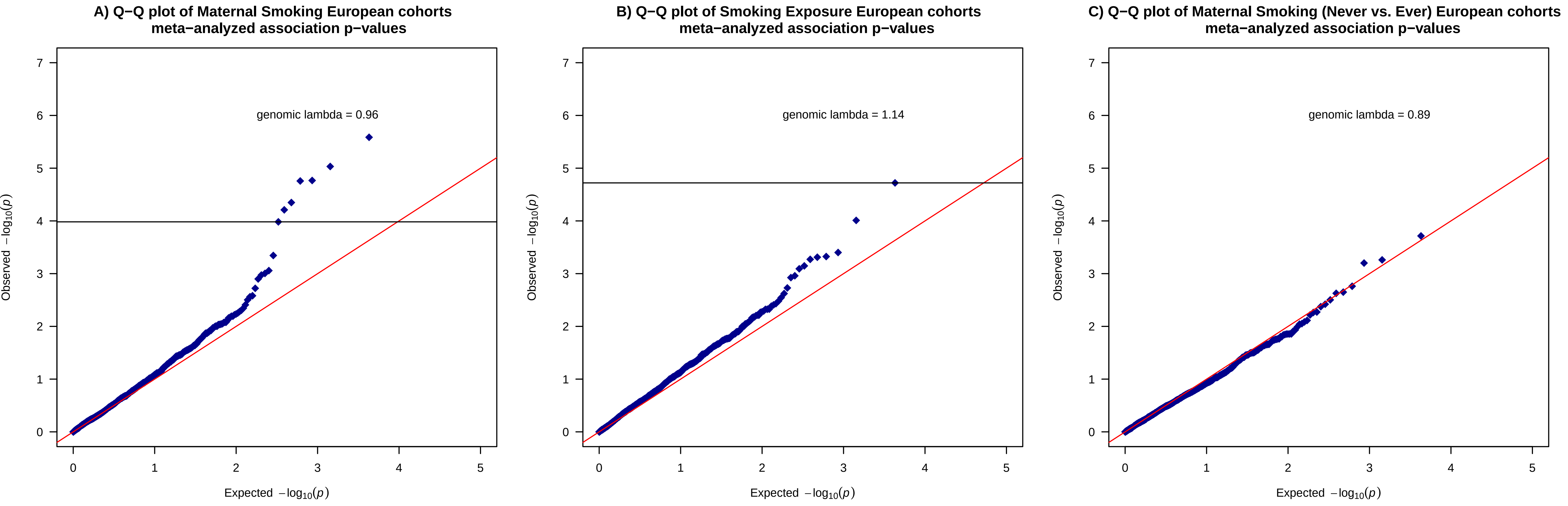

### Supplementary Figure 3

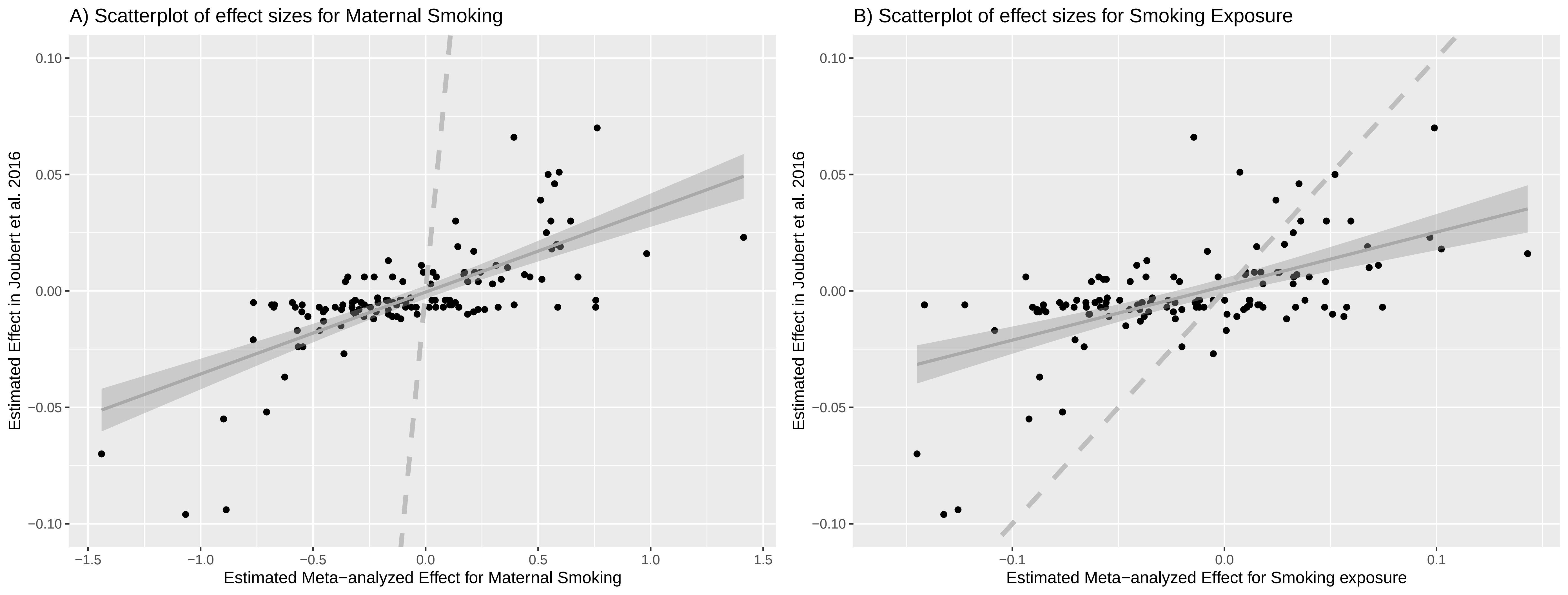

### Supplementary Figure 4

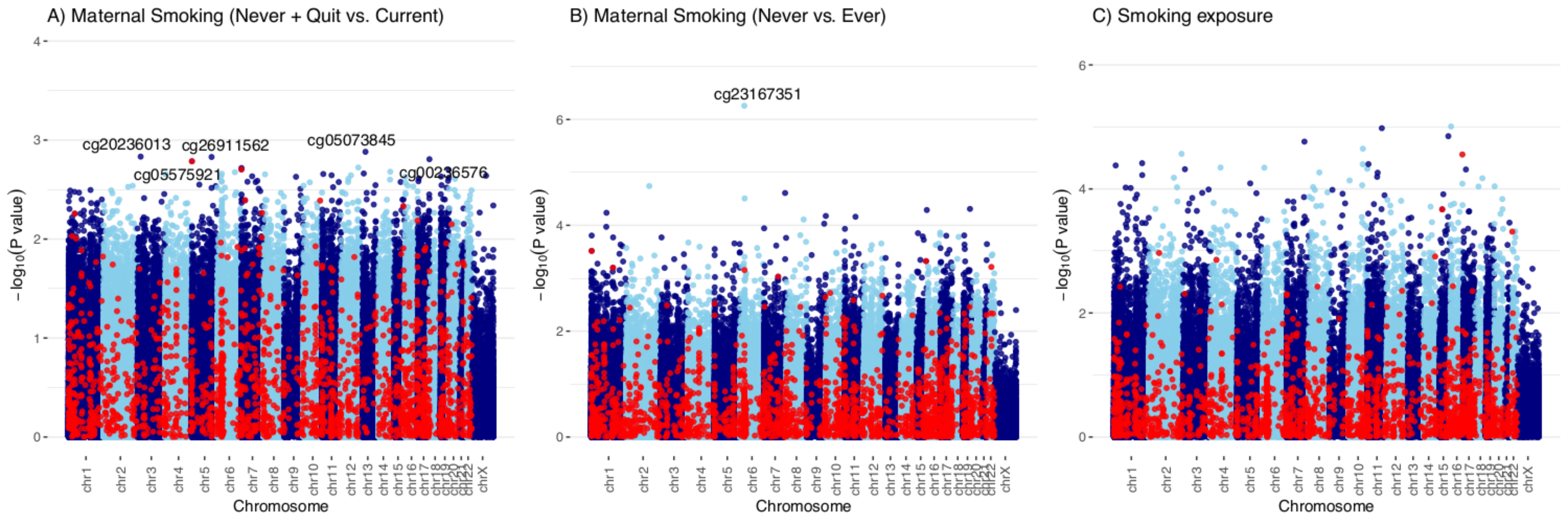

### Supplementary Figure 5

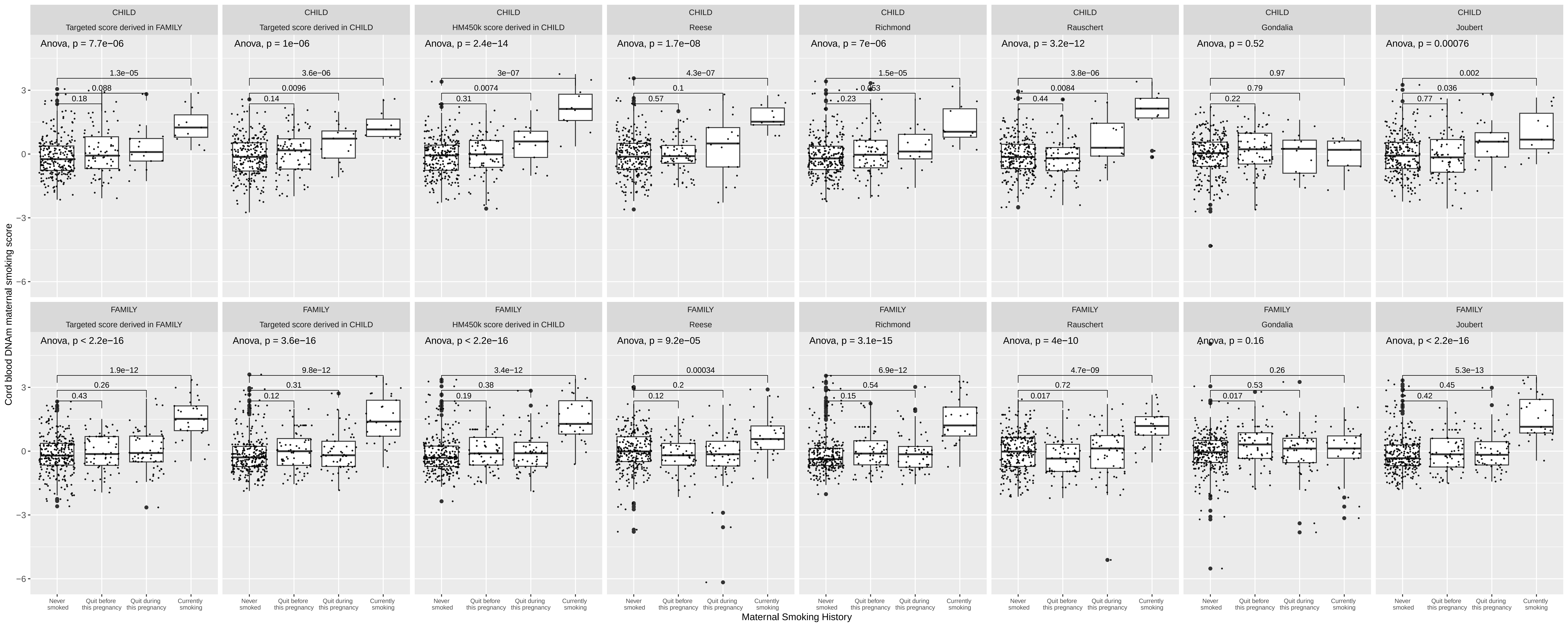

### Supplementary Figure 6

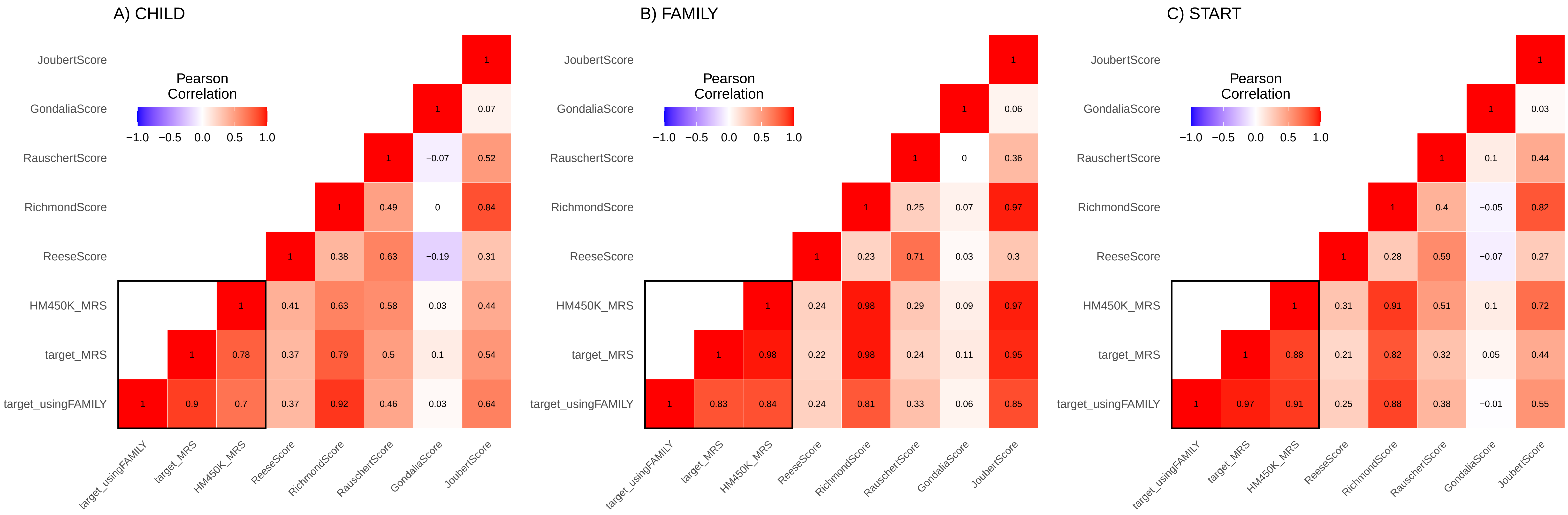

### Supplementary Figure 7

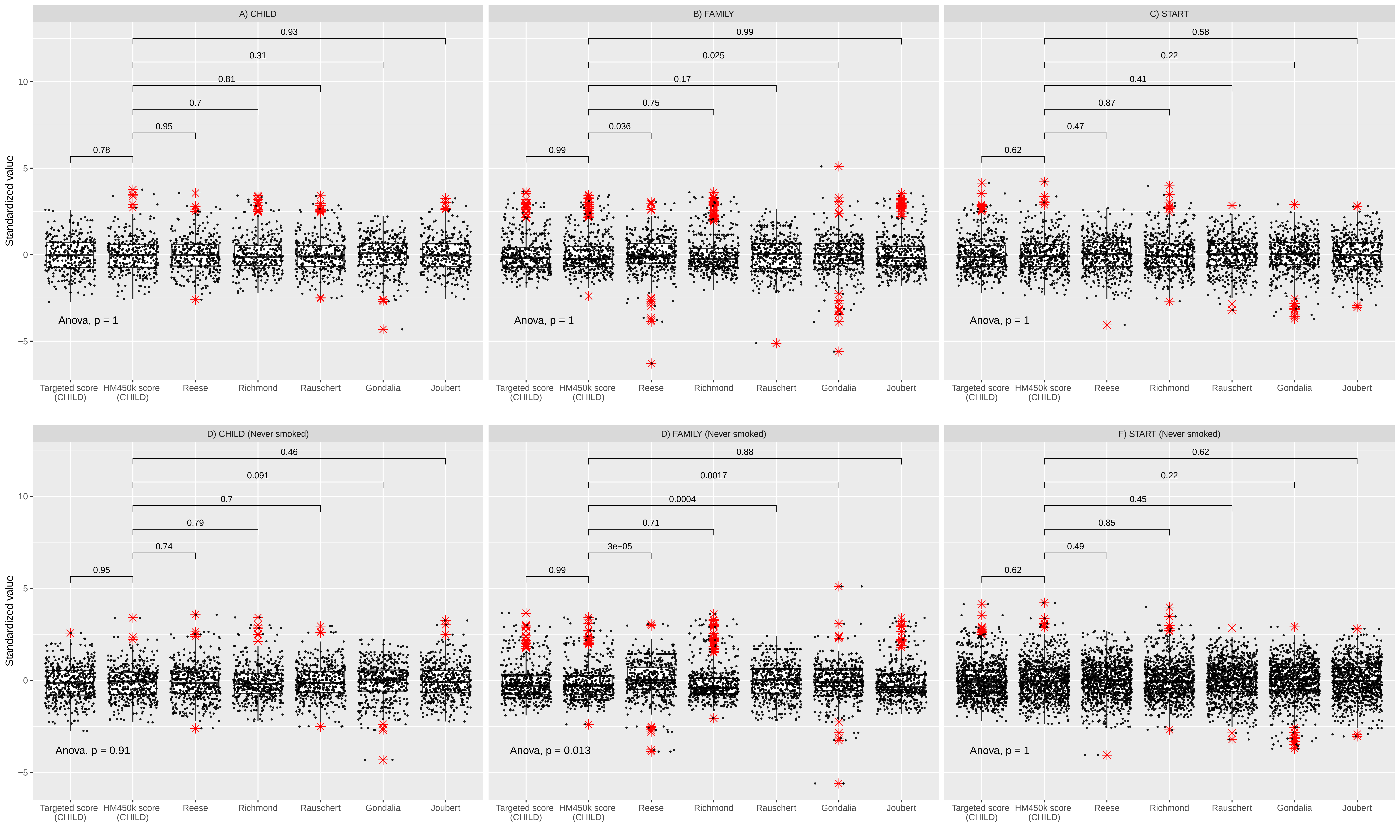
