## Supplementary Material for "Maternal smoking DNA methylation risk score associated with health outcomes in offspring of European and South Asian ancestry"

#### 1 Cord blood DNA methylation (DNAm) data processing and quality controls

Cord blood samples from CHILD and part of START were processed using the Illumina Infinium HumanMethylation450 (HM450) BeadChip; while samples from FAMILY and the remaining START were processed using a custom array designed off of Infinium MethylationEPIC. Each set of data underwent preprocessing and quality control steps using the SeSAMe package [1].

First, raw signal data were processed into $\beta$-values using the standard openSesame work flow outlined (`https://www.bioconductor.org/packages/devel/bioc/vignettes/sesame/inst/doc/sesame.html#The_openSesame_Pipeline`). The function starts by implementing the pOOBAH algorithm for experiment-dependent masking of probes based on signal detection $p$-values. The algorithm uses Infinium-I probe out-of-band (OOB) signal for calibrating the distribution of signal background. Afterwards, "noob" normalization [2] was performed for background subtraction based on normal-exponential deconvolution using the OOB probes. Then, non-linear dye bias correction was applied to equalize the mean of probes from the two color channels. Finally, the corrected probe values were converted into methylation $\beta$-values by taking corrected signal from the methylated allele, and dividing over the sum of methylation and unmethylation corrected signals. The pipeline returned a $\beta$-value matrix for further processing.

In the second step, we applied additional quality control filters to remove samples with $> 10\%$ missing probes and CpG probes with $>$10% samples missing. Further, cross-reactive probes and SNP probes were removed as recommended for HM450 [3] and EPIC arrays [4, 5]. For CpG probes with missing rate $<$10%, mean imputation was performed to fill in the missing values. We further excluded samples that were either mismatches between reported sex and methylation-inferred sex or were duplicates. Finally, we removed non-informative probes that were either all hypomethylated (beta-value < 0.1) or hypermethylated (beta-value > 0.9). These probes might be fine to retain when examined with a symmetric phenotype of interest. But since both the smoking exposure and number of smokers are low in our data, inclusion of these probes could led to a larger than expected number of spurious associations and thus inflated type I error. Removal of these probes have also been shown to have minimal impact on the predictive performance [6].

DNA methylation patterns are known to vary between different tissues and cell types, and meanwhile, many diseases or complex phenotype can also be subjected to cellular heterogeneity, where different cell types coexist within the same tissue. Thus, cell convolution analysis allows us to distinguish changes in DNAm that are specific to certain cell types, and thus control potential bias that arise from the association between DNAm signals at individual CpGs and phenotypes of interest. A joint umbilical cord blood reference library constructed from four reference datasets [7] was used for deconvolution in our cord blood samples. The cell-type proportions were estimated for CD8T, CD4T, Natural Killer cells, B cells, monocytes, granulocytes, and nucleated red blood cells.

#### 2 Statistical Notation and Methods

Let $y \in \mathcal{R}^n$ denote a continuous outcome of interest, such as smoking exposure, that has been mean and variance standardized. We hope to establish a linear regression model that can explain the variation in $y$ using linear combinations of $X$:

$$y = X\gamma + \epsilon, \tag{2.1}$$

where $X \in \mathcal{R}^{n\times p}$ denotes a column standardized $\beta$-matrix of the $p$ CpGs measured on $n$ individuals. But many of the CpGs in physical proximity are often correlated, causing instability in converging to a solution and/or leading to variance inflation in the resulting coefficients when estimated simultaneously.

A lasso solution [8] was designed to alleviate the multi-collinearity of this estimation problem and can be obtained by minimizing the objective function that includes an L-1 penalty term that regularizes $\gamma$, forcing some of the coefficients to be exactly zero:

$$\hat{\gamma} = \min_{\gamma} \Big\{ (y - X\gamma)^T (y - X\gamma) + 2\lambda \sum_{j=1}^{p} |\gamma_i| \Big\}. \tag{2.2}$$

This can be done directly in a dataset with individual level data $X$ and $y$ of sample size $n$, however, this approach is potentially underpowered for testing dataset with small sample size as is the case here. We proposed to adopt the approach outlined in [9], establishing an equivalent optimization solution under the lasso constraint but using the summary statistics from a larger discovery samples (sample size $N > n$).

We modified their elastic net solution that depended on two tuning parameters, as well as inputs, namely the summary statistics and a reference CpG data covariance matrix. The Elastic net using summary statistics function is available from the R package "lassosum" (`https://github.com/tshmak/lassosum/blob/0e44b530c77e862f1b88f9e1fad88fdab44e7827/R/elnetR.R`), where both the L-1 and L-2 penalty parameters $\lambda_1$ and $\lambda_2$ needed to be selected.

To select the optimal tuning parameters, we examined a range of $\lambda_1$ values that forces all weights to be zero or no penalty, with 50 incremental increases, and $lambda_2$ was taken to be $\alpha(1 - \lambda_1)$ where $\alpha$ was set to be 0 to 1 with incremental increases of 0.1. These together gave a grid of $10 \times 50$ choices for the two tuning parameter values. The tuning parameter pair that produced a score that was most significantly associated with the smoking history variable, without any data transformation, was chosen as final elastic net solution.

### 3 Sensitivity analyses

#### 3.1 Sensitivity to genetic ancestry

To examine the impact of not restricting to genetically confirmed samples, here we test for differences in the beta-values of the top associated CpGs for maternal smoking and the derived MRS between samples with genetically confirmed vs. no or ambiguous genetic ancestry. For each study, the genetic ancestry was called by examining the top 2 genetic principal components from the combined cohort and 1000 Genome Project metropolitan reference panel [10] phase 3 release [11]. Obvious outliers in both x- and y-axes were removed. Those that did not fall within 95% confidence region of the cluster center as defined by the metropolitan populations of 1000 Genomes samples were flagged as potentially ambiguous ancestry, otherwise each sample was assigned an ancestry based on their proximity to the 1000 Genome sample ancestry groupings. Any participant that did not provide self-reported ancestry or had no genetic data had already been removed as per our quality control protocol (Supplementary Table 1).

We compared the difference in distribution of methylation levels or scores between the genetically confirmed group vs. others using a two-sample Kolmogorov–Smirnov test, which is a non-parametric test without distribution assumptions, due to the distribution of the cpgs being non-normal and between 0 and 1. The p-value for each cpg, MRS was tabulated below. The results suggested no material difference in the distribution between those that were genetically confirmed Europeans (or South Asians) without those that self-reported but not genetically confirmed. Of note, there was marginal evidence of the Reese and Rauschert scores to be slightly different in FAMILY, but not in CHILD nor START.

#### 3.2 Smoking exposure regression model diagnostics

Smoking exposure, measured by the number of hours exposured to cigarette smoking per week, was highly skewed and zero-inflated across the three cohorts, as shown in Figure 1 below.

| | CHILD | FAMILY | START |
|---|---|---|---|
| full samples (n) | 352 | 411 | 504 |
| genetically confirmed samples (n) | 268 | 312 | 488 |
| cg12876356 | 0.72 | 0.80 | 0.77 |
| cg09935388 | 0.48 | 0.33 | 0.55 |
| cg14179389 | 0.94 | 0.72 | 0.30 |
| cg18146737 | 0.67 | 0.22 | 0.29 |
| cg09662411 | 0.84 | 0.34 | 0.81 |
| cg18316974 | 0.73 | 0.68 | 0.41 |
| cg01798813 | 0.23 | 0.35 | 0.42 |
| Reese Score | 0.30 | 0.01 | 0.90 |
| Richmond Score | 0.48 | 0.99 | 0.58 |
| Rauschert Score | 0.23 | 0.02 | 0.10 |
| Gondalia Score | 0.64 | 0.18 | 0.71 |
| Jourbet Score | 0.37 | 0.94 | 0.67 |
| HM450K Score | 0.49 | 0.83 | 0.87 |
| target Score | 0.22 | 0.97 | 0.29 |

Table 1: Mean difference in methylation level and scores between studies

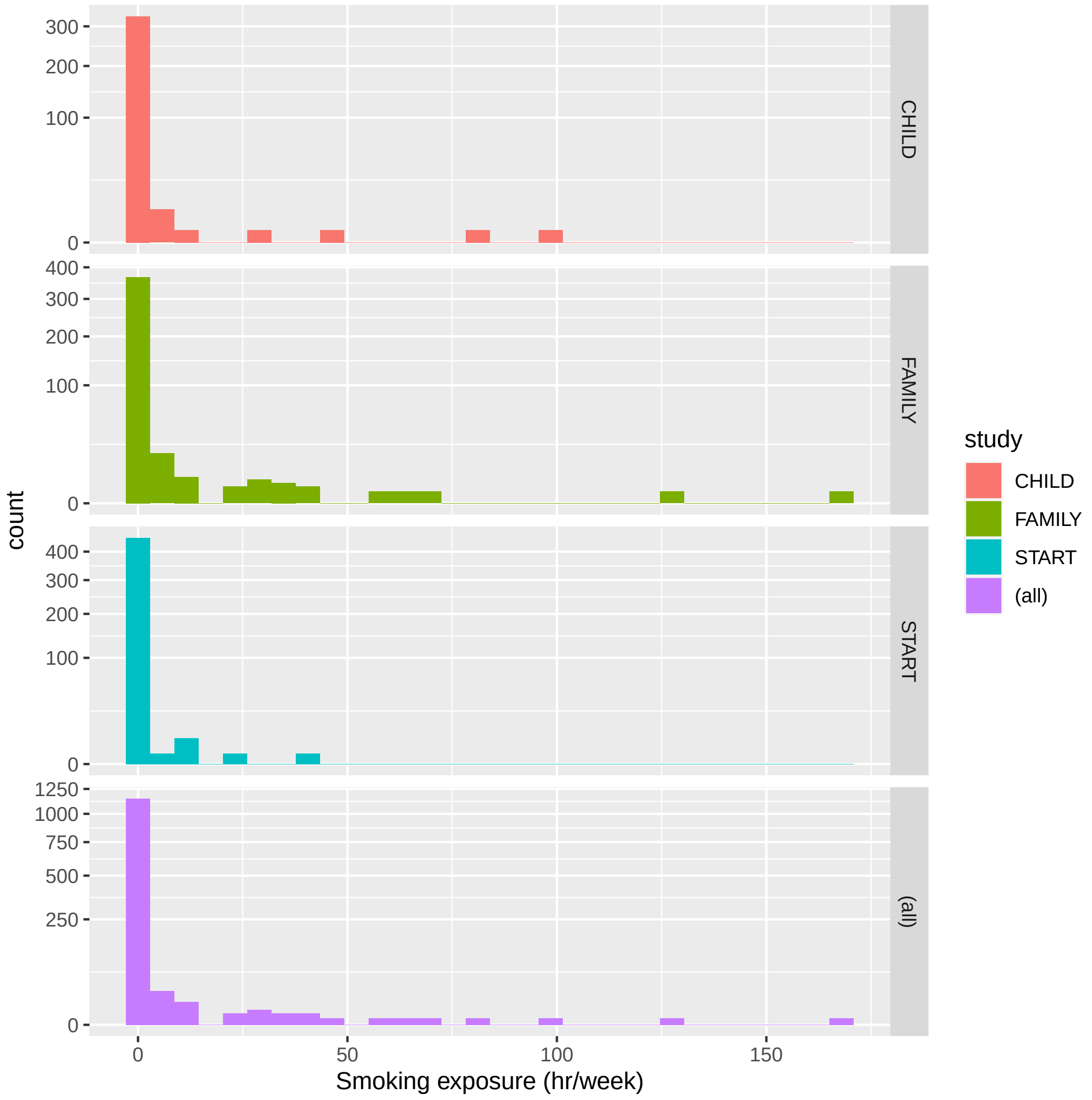


Figure 1: Histogram of the smoking exposure across the three cohorts.

Figures 2 and 4 showed the model diagnostics using CHILD and FAMILY data, where the departure from linearity (measured by distance from the blue line to each point) was quite severe. There were also considerable variance heteroskedasticity as shown in the scale and location diagnostic plot. In both data, the main outlying points corresponded to the tail of the smoking exposure phenotype (>= 25 hr/week).

Figures 3 and 5 showed the improved model diagnostics using CHILD and FAMILY data after data transformation. In particular, the normality assumption had improved greatly, along with reduced distance in terms of departure from constant variance, linearity, and outlying values.

The association results below also suggested potentially better control of type I error rates as marked by the reduced p-values.

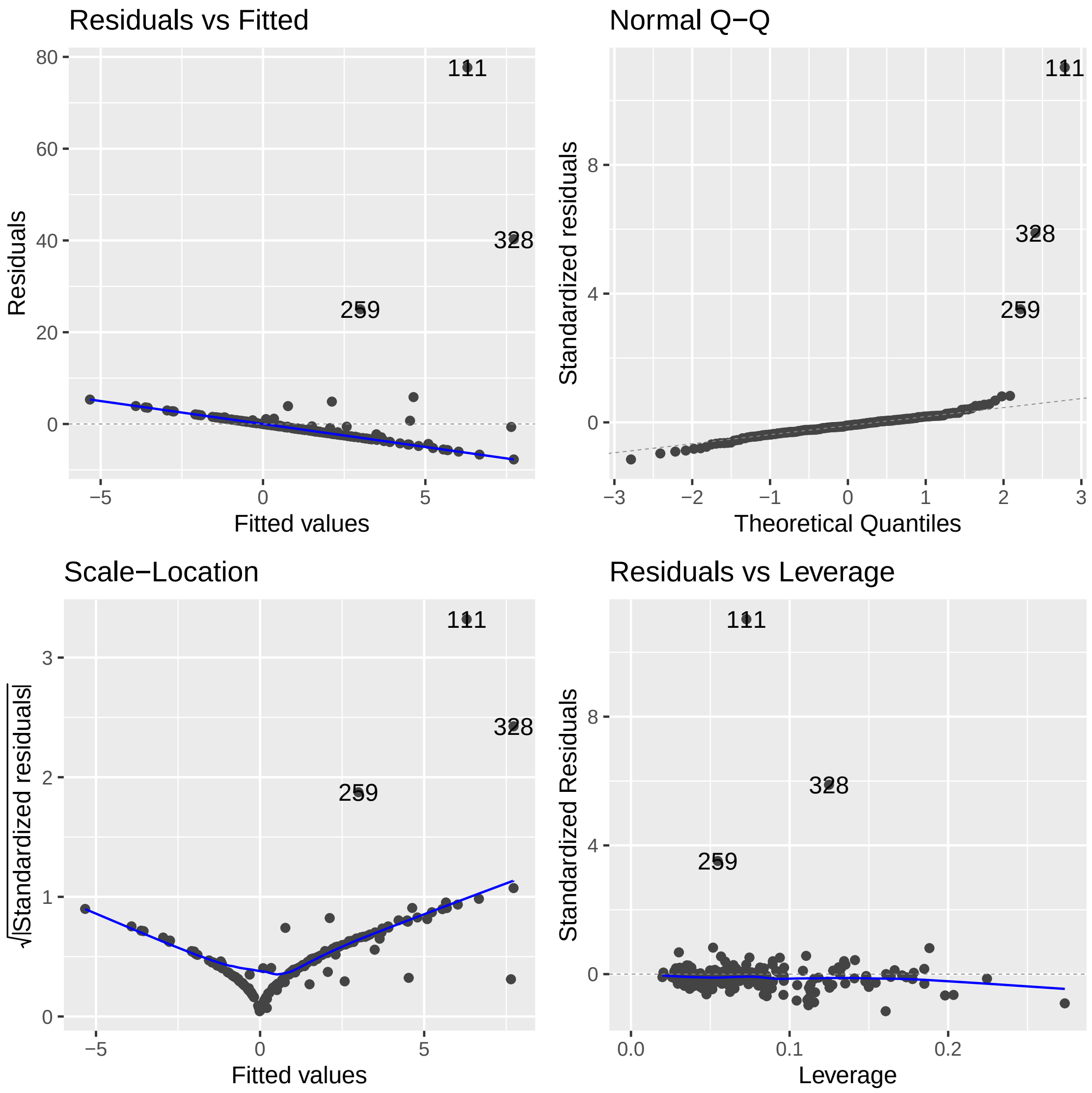


Figure 2: Diagnostic for smoking exposure (raw) in CHILD.

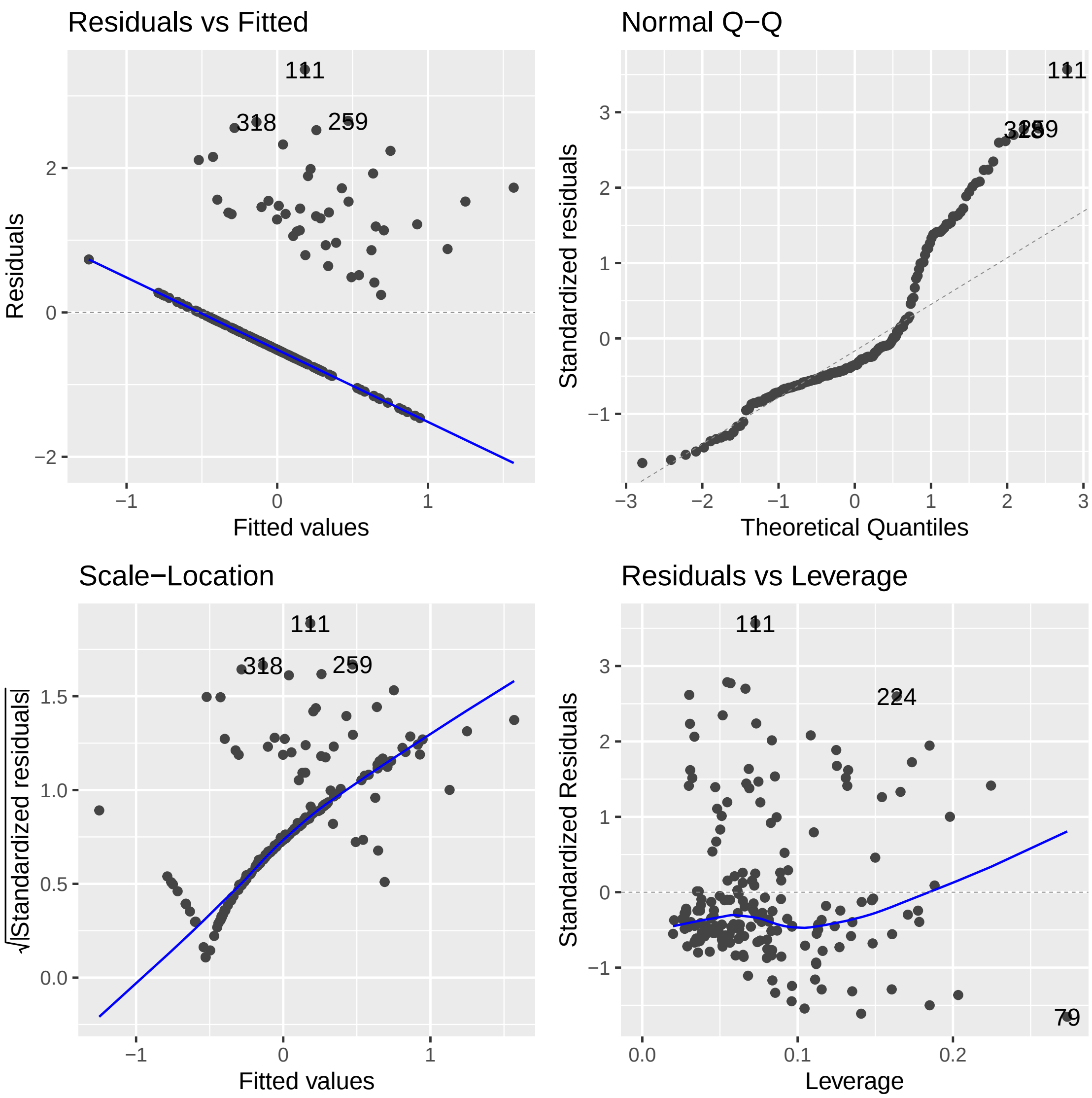


Figure 3: Diagnostic for smoking exposure (rank-transformed) in CHILD.

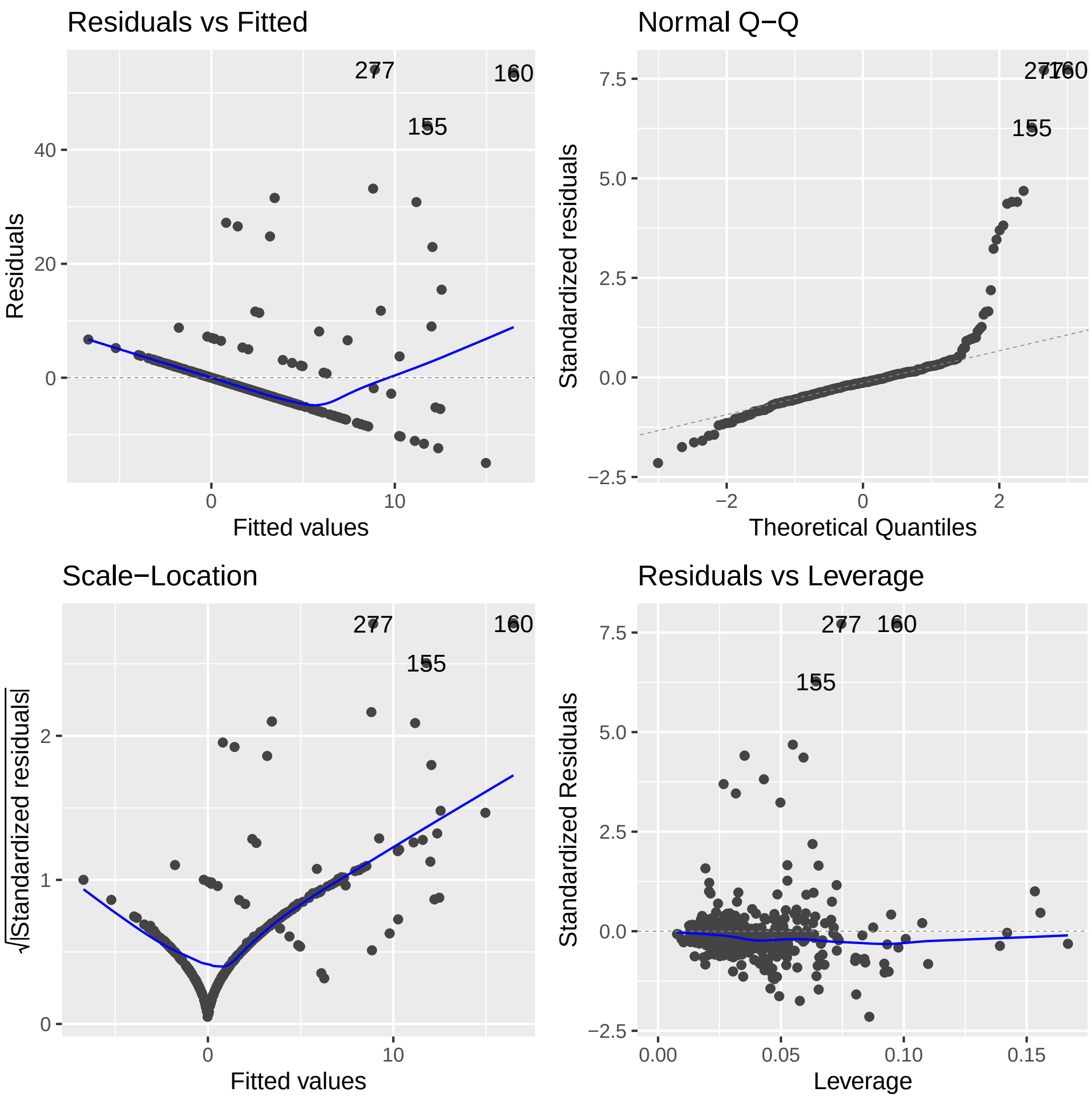


Figure 4: Diagnostic for smoking exposure (raw) in FAMILY.

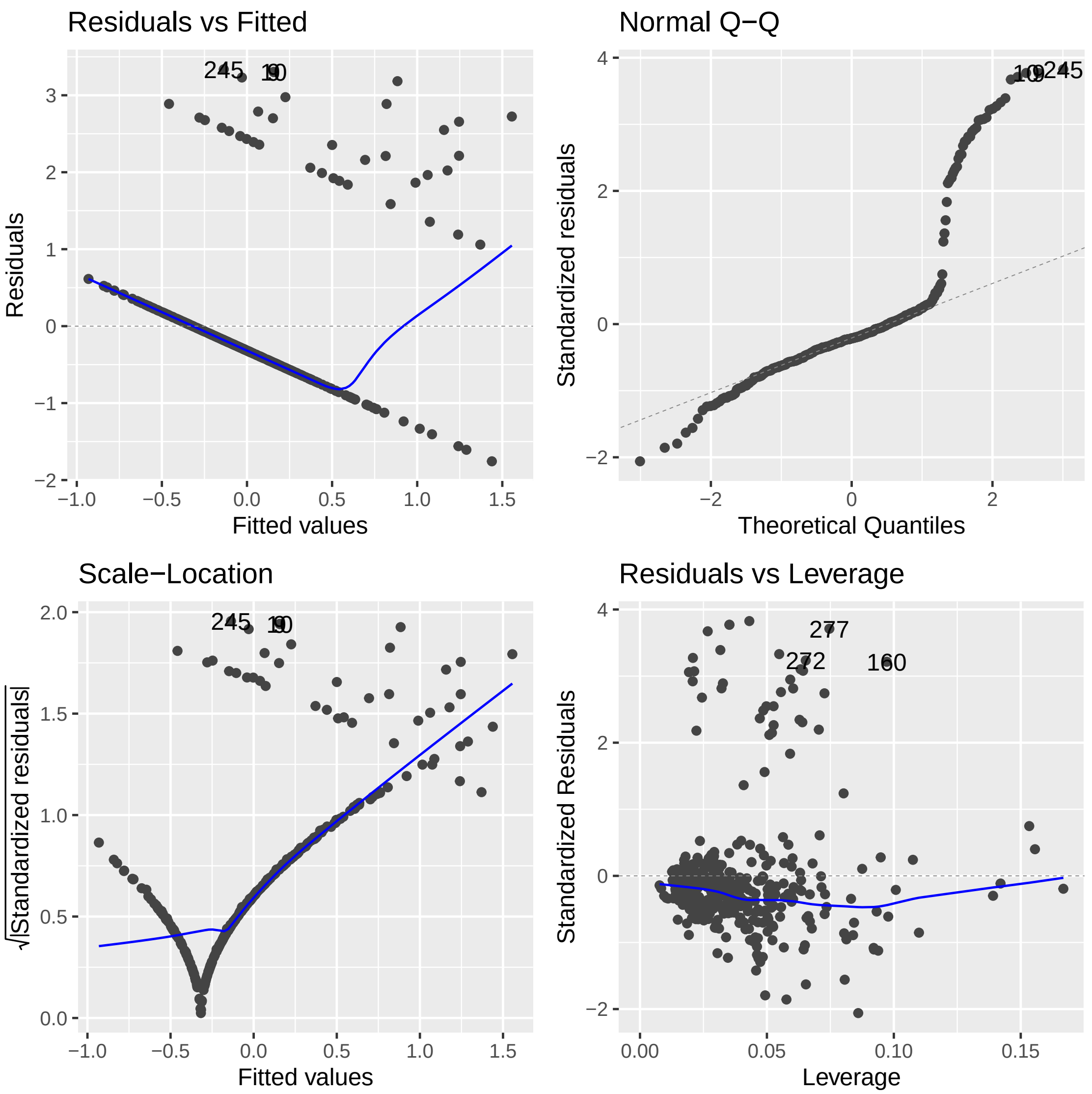


Figure 5: Diagnostic for smoking exposure (rank-transformed) in FAMILY.

```
lm(formula = as.formula(paste(paste("mblsmkexp", "~"), "cg09935388",
    " + agemom + parity + ppbmi + mblsdi + mbleduyrs + GDM + CD8T + CD4T + NK +
   Bcell + Mono + Gran + nRBC")),
    data = analytical_CHILD450)

Residuals:
   Min     1Q Median     3Q    Max
-7.723 -1.988 -0.676  0.623 77.705

Coefficients:
            Estimate Std. Error t value Pr(>|t|)
(Intercept) -85.7152    47.2971  -1.812   0.0717 .
cg09935388   -8.0699     4.6175  -1.748   0.0823 .
agemom       -0.1318     0.1484  -0.888   0.3759
parity        1.3020     0.7514   1.733   0.0849 .
ppbmi         0.1298     0.1121   1.157   0.2488
mblsdi       -0.5836     0.6499  -0.898   0.3704
mbleduyrs    -0.2320     0.2006  -1.156   0.2491
GDM          -1.6081     2.3699  -0.679   0.4983
CD8T         71.2223    55.7042   1.279   0.2028
CD4T         95.9692    50.9789   1.883   0.0614 .
NK           75.4159    48.2076   1.564   0.1196
Bcell        98.5986    49.4326   1.995   0.0477 *
Mono        113.5933    60.8135   1.868   0.0635 .
Gran         92.6992    45.7433   2.027   0.0442 *
nRBC         89.3165    45.0392   1.983   0.0489 *
---

Residual standard error: 7.318 on 173 degrees of freedom
  (164 observations deleted due to missingness)
Multiple R-squared:  0.09055, Adjusted R-squared:  0.01695
F-statistic:  1.23 on 14 and 173 DF,  p-value: 0.2572

lm(formula = as.formula(paste(paste("mblsmkexp_inv", "~"), "cg09935388",
    " + agemom + parity + ppbmi + mblsdi + mbleduyrs + GDM + CD8T + CD4T + NK +
   Bcell + Mono + Gran + nRBC")),
    data = analytical_CHILD450)

Residuals:
    Min      1Q  Median      3Q     Max
-1.4633 -0.5538 -0.3354  0.2358  3.3643

Coefficients:
              Estimate Std. Error t value Pr(>|t|)
(Intercept) -17.751016   6.331899  -2.803  0.00563 **
cg09935388   -0.937767   0.618173  -1.517  0.13109
agemom       -0.004649   0.019872  -0.234  0.81529
parity        0.077288   0.100593   0.768  0.44334
ppbmi         0.017526   0.015012   1.167  0.24463
mblsdi        0.080070   0.087001   0.920  0.35868
mbleduyrs    -0.060883   0.026858  -2.267  0.02464 *
GDM          -0.076305   0.317274  -0.241  0.81022
CD8T         22.823937   7.457404   3.061  0.00256 **
CD4T         17.575277   6.824799   2.575  0.01085 *
NK           20.063766   6.453790   3.109  0.00220 **
Bcell        17.137434   6.617786   2.590  0.01043 *
Mono         16.010166   8.141417   1.967  0.05084 .
Gran         18.200666   6.123886   2.972  0.00338 **
nRBC         18.561780   6.029630   3.078  0.00242 **
---

Residual standard error: 0.9797 on 173 degrees of freedom
  (164 observations deleted due to missingness)
Multiple R-squared:  0.1645,  Adjusted R-squared:  0.09693
F-statistic: 2.434 on 14 and 173 DF,  p-value: 0.003807
```

Listing 1: R output for CHILD

```
lm(formula = as.formula(paste(paste("mblsmkexpng", "~"), "cg09935388",
    " + agemom + parity + ppbmi + mblsdi + mbleduyrs + GDM + CD8T + CD4T + NK +
    Bcell + Mono + Gran + nRBC")),
    data = analytical_FAMILY)

Residuals:
    Min      1Q  Median      3Q     Max
-14.965  -2.841  -0.917   0.992  54.084

Coefficients:
             Estimate Std. Error t value Pr(>|t|)
(Intercept)   9.42811   19.49932   0.484   0.6290
cg09935388  -13.21789    3.26557  -4.048 6.32e-05 ***
agemom       -0.16439    0.08384  -1.961   0.0507 .
parity       -0.10878    0.43349  -0.251   0.8020
ppbmi         0.05438    0.06550   0.830   0.4070
mblsdi        1.51775    0.32423   4.681 4.03e-06 ***
mbleduyrs    -0.11365    0.12490  -0.910   0.3635
GDM           1.47034    1.11635   1.317   0.1886
CD8T         18.24084   25.50787   0.715   0.4750
CD4T          9.98686   20.78984   0.480   0.6313
NK            9.48095   24.27533   0.391   0.6964
Bcell       -14.75148   23.73408  -0.622   0.5346
Mono         24.62385   22.68272   1.086   0.2784
Gran          6.24709   19.33151   0.323   0.7468
nRBC         -1.51381   19.55469  -0.077   0.9383
---

Residual standard error: 7.286 on 364 degrees of freedom
  (32 observations deleted due to missingness)
Multiple R-squared:  0.1843,  Adjusted R-squared:  0.1529
F-statistic: 5.873 on 14 and 364 DF,  p-value: 2.055e-10


lm(formula = as.formula(paste(paste("mblsmkexp_inv", "~"), "cg09935388",
    " + agemom + parity + ppbmi + mblsdi + mbleduyrs + GDM + CD8T + CD4T + NK +
    Bcell + Mono + Gran + nRBC")),
    data = analytical_FAMILY)

Residuals:
    Min      1Q  Median      3Q     Max
-1.7566 -0.4259 -0.1868  0.0594  3.3374

Coefficients:
             Estimate Std. Error t value Pr(>|t|)
(Intercept)  0.462904   2.386374   0.194 0.846301
cg09935388  -1.442728   0.399648  -3.610 0.000349 ***
agemom      -0.018153   0.010260  -1.769 0.077678 .
parity      -0.042887   0.053051  -0.808 0.419380
ppbmi        0.008628   0.008016   1.076 0.282506
mblsdi       0.176502   0.039680   4.448 1.15e-05 ***
mbleduyrs   -0.029251   0.015286  -1.914 0.056461 .
GDM          0.141265   0.136622   1.034 0.301829
CD8T         1.844482   3.121716   0.591 0.554984
CD4T         2.161755   2.544312   0.850 0.396082
NK           1.506381   2.970874   0.507 0.612427
Bcell       -0.718622   2.904635  -0.247 0.804734
Mono         4.140470   2.775966   1.492 0.136685
Gran         1.173117   2.365838   0.496 0.620295
nRBC         0.250795   2.393151   0.105 0.916595
---

Residual standard error: 0.8917 on 364 degrees of freedom
  (32 observations deleted due to missingness)
Multiple R-squared:  0.1833,  Adjusted R-squared:  0.1519
F-statistic: 5.834 on 14 and 364 DF,  p-value: 2.49e-10
```

Listing 2: R output for FAMILY
